# Clinical pathogen adjudication in metagenomic sequencing using a large language model

**DOI:** 10.64898/2026.10.16.26361411

**Authors:** Muyun Wei, Yiming Zhao, Chao Fang, Shengyang Zhang, Ke Xu, Shuangshuang Li, Lixue Liu, Yanmeng Pang, Lungang Liang, Ziqing Deng, Min Li

**Affiliations:** Department of Laboratory Medicine, Ren Ji Hospital, Shanghai Jiao Tong University School of Medicine, Shanghai 200127, China; BGI Research, Beijing, China; Shanghai Jiao Tong University School of Medicine, Shanghai 200127, China; Shenzhen Key Laboratory of Unknown Pathogen Identification, BGI Research, Shenzhen 518083, China; State Key Laboratory of Genome and Multi-omics Technologies, BGI Research, Shenzhen 518083, China; Clinical Laboratory, BGI Genomics, Shanghai, China

## Abstract

Clinical metagenomic sequencing detects hundreds of organisms whose relevance to infection requires clinicians’ manual interpretation. We retrospectively reviewed 4,855 specimens from 19 sampling sites, linked 1.33 million detected organisms to clinical culprit-pathogen determinations, and fine-tuned a large language model on these labels. The model recovered 92.44% of reference culprit calls in the held-out test set and 84.49% in a later cohort. In a blinded re-adjudication of 215 cases, the model matched 90.5% of culprit calls on which both expert reviewers agreed. Adjudication accuracy varied with organism identity and specimen types. mNGS-derived metrics such as read counts or genome coverage alone were insufficient to identify culprit pathogens, and organisms classified as common colonizers could still be judged causative. Certain organisms were frequently co-reported as causative in the same specimen. These findings support further evaluation of human and AI-assisted report review, particularly regarding missed pathogens, excess calls, and differences across specimen types.

## Introduction

Metagenomic next-generation sequencing (mNGS) has entered clinical use in the evaluation of severe or diagnostically unresolved suspected infection^1–4^. Clinical assays have been deployed in cerebrospinal fluid, plasma and respiratory specimens, extending the range of organisms that can be investigated from a single sample^1,4–6^. This breadth also creates an interpretation problem. A report may contain hundreds of organisms, most of which represent colonisation, environmental or reagent background, or detections of uncertain clinical relevance^7,8^. The clinically decisive step is to determine which detected organisms could explain the patient’s current infection. Both overlooking a relevant pathogen and overinterpreting an incidental finding can affect the subsequent diagnostic assessment.

This judgement is usually made by clinical microbiologists and treating teams who read the report alongside the patient’s clinical record. Quantitative indices, including read counts, relative abundance and genome coverage, provide evidence for interpretation, while organism identity, specimen type and clinical context affect what that evidence means. Reporting recommendations and decision-support approaches commonly use quantitative thresholds and curated organism annotations^9–11^. However, the information supporting clinical adjudication has been difficult to examine systematically. Much computational work in mNGS addresses read classification, host depletion, contamination and abundance estimation^12–14^. Resources that connect complete detection panels to the treating team’s assessment of causative relevance are needed to evaluate the subsequent interpretation step.

Large language models can integrate structured and narrative information and have been evaluated on clinical knowledge and diagnostic reasoning tasks^15–18^. Their performance on medical questions or clinical-document benchmarks does not establish their usefulness for interpreting the many competing candidates in an mNGS report^19,20^. Recent studies have also applied language models to metagenomic report interpretation^21,22^. These efforts are constrained by a more fundamental condition: what limits AI-assisted mNGS interpretation is no longer data or compute, but the absence of large, real-world cohorts with high-quality labels, clinical endpoints and clinical information. Without such cohorts, previous evaluations could not adequately address several key dimensions: how reproducible the reference judgement itself is, the recovery of relevant candidates alongside additional positive calls, specimen-type differences, disagreement among clinical readers, and why additional contextual information fails to improve predictions.

Here, we assembled an expert-labelled resource of 4,855 mNGS specimens spanning 19 specimen types and 1.33 million organism detections. We defined culprit-pathogen adjudication as the prediction of the treating team’s causative call for each organism in a specimen, evaluated a fine-tuned language model against this reference, and compared its predictions with an independent blinded re-adjudication. We then examined how quantitative evidence, species identity, specimen source and co-adjudicated organisms relate to the decision. The model serves both as a reference system for the task and as an analytical instrument for studying its difficulties. Our aim was to identify the capabilities and limitations that matter when considering automated support for clinical report review.

## Results

### Large, expert-labelled mNGS cohorts linking complete detection panels to clinical culprit-pathogen judgements

We assembled 4,855 specimens from two cohorts. The earlier cohort comprised 3,693 specimens tested in 2024 or earlier, of which 2,479 were randomly selected for development and 1,214 were held out; the later cohort comprised 1,162 specimens tested in the first quarter of 2025 (Fig. 1a,b). Each candidate record represents one reported organism in one specimen and contains sequencing indices, report annotations, sampling site and available clinical information. The reference label is the treating team’s recorded causative-pathogen judgement. The corpus contains 1,333,867 candidate records.

**Figure 1.**
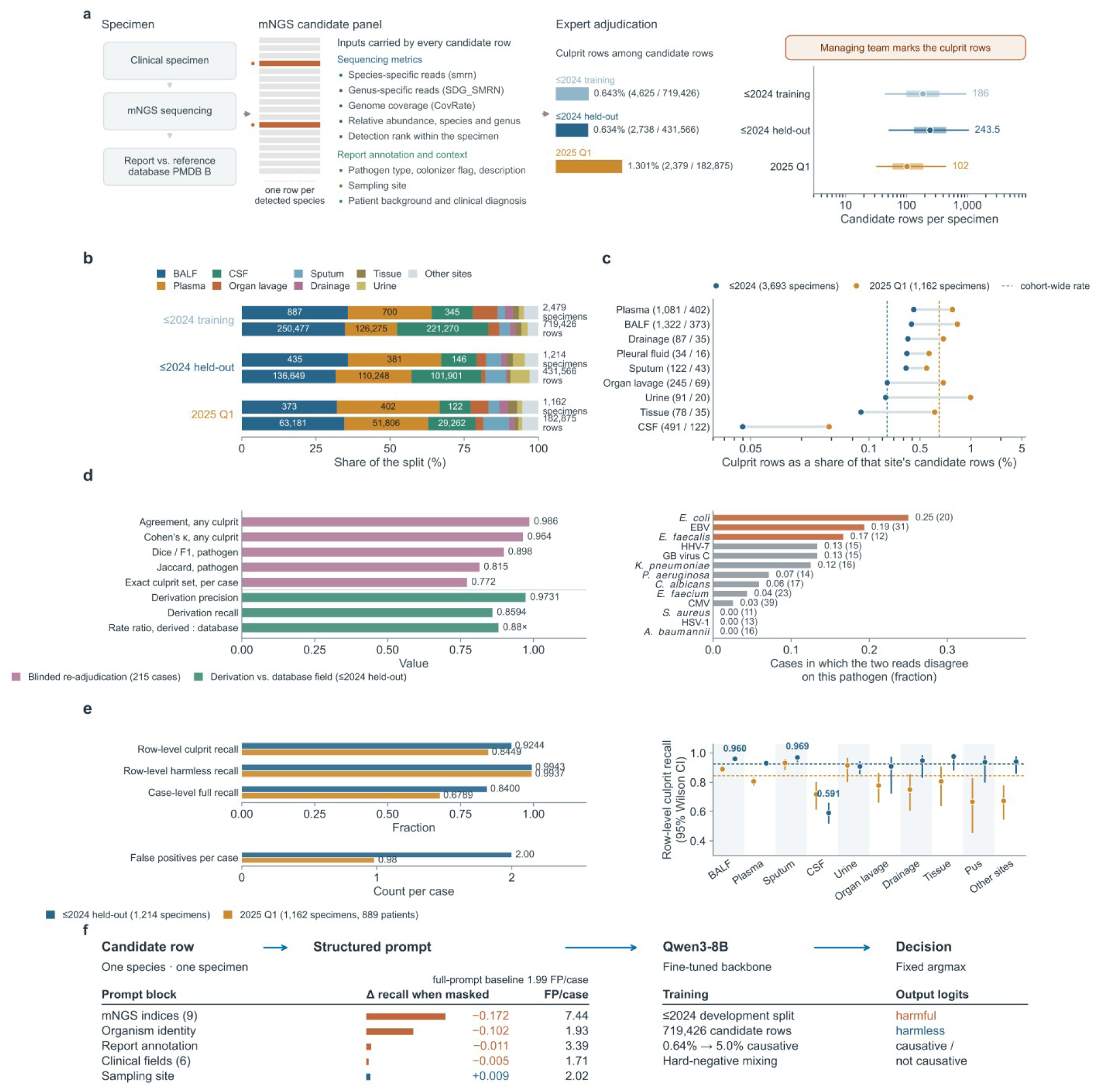
Clinical cohort and evaluation of culprit pathogen adjudication. **a**, The adjudication task: each specimen yields a candidate panel of one row per detected species, carrying sequencing metrics, the report annotation, the sampling site and the clinical information, and the managing team marks the causative rows. Bars, causative rows as a share of all candidate rows in each split. Right, candidate-panel size per specimen: dot, median; bar, interquartile range; line, 5th–95th percentile (n = 2,479, 1,214 and 1,162 specimens). **b**, Corpus composition by sampling site; per split the upper bar is the specimens and the lower bar the candidate rows, the eight commonest sites separate and the rest pooled. Numbers in segments are counts, totals at the right. **c**, Causative rows as a share of that site’s candidate rows, log scale, for the pooled ≤2024 cohort (blue) and 2025 Q1 (orange); parentheses, specimen counts; dashed lines, cohort-wide rates. **d**, Label-extraction checks and blinded clinical re-adjudication: green, reconstructed labels compared with an independent database field on the ≤2024 held-out cohort; purple, agreement between the original and blinded clinical assessments of 215 specimens. Right, the fraction of cases in which the two assessments disagree on a pathogen. **e**, Model baselines. Left, three rate metrics for both cohorts on a common 0–1 axis, false positives per case on its own axis below. Right, causative-row recall by site with 95% Wilson intervals; dashed lines, cohort-wide recalls. **f**, Technical route, and the change in causative-row recall when each prompt block alone is masked (bar length, |Δ recall|; vermilion, worse than the full-prompt baseline of 1.99 false positives per case; blue, better). Inclusion thresholds and model settings are in Methods.

For analyses of species frequency and co-adjudication, reports were normalised to avoid counting the same organism more than once within a specimen. Merging multi-granularity reports, species complexes and renamed taxa reduced the culprit counts by 11.7% in the earlier held-out cohort and 8.4% in 2025 Q1 (Supplementary Fig. 1). This normalization concerns reporting units within a specimen; separate specimens remain separate testing events.

Only a small proportion of organism detections were judged causative, and this proportion differed by specimen source. In the earlier cohort, causative labels accounted for 0.919% of candidate records in plasma and 0.090% in cerebrospinal fluid (CSF; Fig. 1c). Among the 2,376 specimens in the held-out and 2025 Q1 cohorts, 49.1% had at least two organisms judged causative, whereas 23.4% had no organism judged causative. Clinical interpretation therefore involved determining which, if any, of the organisms detected in a specimen could explain the current infection.

### A fine-tuned language model for culprit-pathogen adjudication, and its performance in held-out and recent cohorts

We fine-tuned Qwen3-8B^23^ on the development cohort and evaluated it with a fixed binary decision rule (Fig. 1e,f). Recall at the record level was 92.44% in the earlier held-out cohort and 84.49% in 2025 Q1, with 2.00 and 0.98 false-positive records per specimen, respectively. The model recovered all reference culprits in 84.00% and 67.89% of culprit-bearing specimens. This measure tolerates additional positive predictions and does not imply a fully correct report.

Performance also differed by specimen source. In the earlier held-out cohort, culprit recall was 96.0% in bronchoalveolar lavage fluid (BALF), 93.0% in plasma and 59.1% in CSF; in the later cohort, the corresponding values were 89.0%, 80.6% and 71.8%.

Sixteen baseline models were evaluated on the same candidate records and labels. A gradient-boosted tree model using quantitative indices, organism identity, sampling site and report text achieved an average precision of 0.707. When fitted to the same culprit-enriched training data as the language model, its average precision increased to 0.798, compared with 0.825 for the language model. At their respective operating points, the language model recovered 252 more reference culprits than the baseline in the earlier test cohort but generated 790 more false positives, which means 3.1 additional false positives per additional culprit recovered. The corresponding ratio was 1.4 in 2025 Q1. The comparison therefore identifies a recall–false-positive trade-off whose clinical value requires evaluation in the intended review workflow.

### Interpretation of sequencing evidence depends on organism identity, specimen source and combined indices, not on any single index

We next examined the information associated with clinical adjudication. Within 168 species–site combinations containing at least five culprit and five non-culprit detections, several individual sequencing indices separated the two classes well (Fig. 2c). This analysis used the pooled earlier cohort (development and held-out specimens) and 61 qualifying combinations in 2025 Q1. Median area under the receiver-operating characteristic curve (AUC) was 0.922 for genome coverage, 0.913 for species relative abundance and 0.869 for species-specific reads. Nevertheless, the best single index reached an average precision of only 0.595 in the earlier cohort and 0.584 in the later cohort. Combining five indices increased average precision to 0.741 and 0.750, respectively (Fig. 2d). These estimates are based on selected species–site combinations, not the complete candidate panels used for the model comparison. Though quantitative evidence was informative, single index was insufficient to reproduce the clinical adjudication.

**Figure 2.**
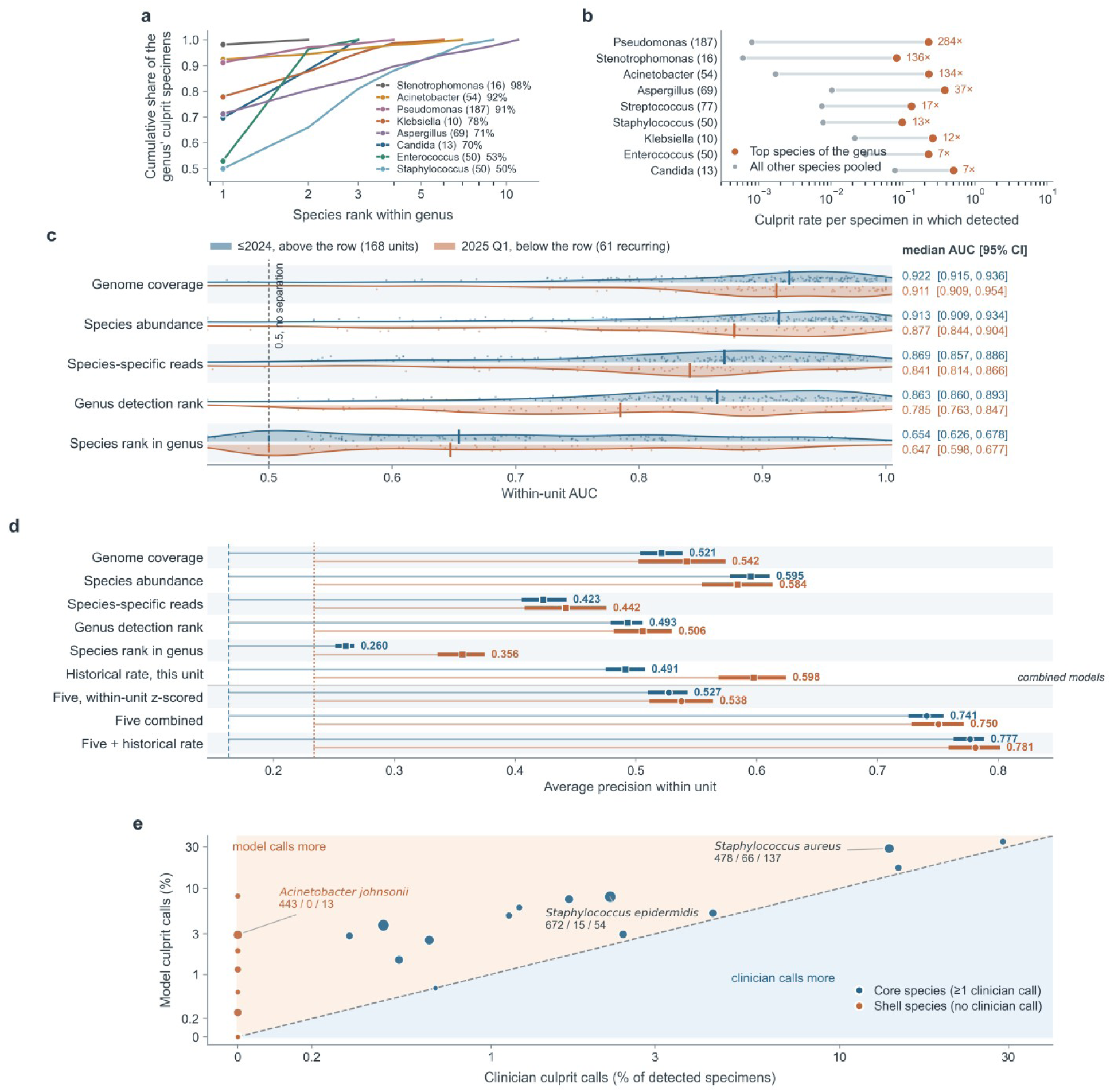
Quantitative sequencing evidence and species identity in clinical adjudication. a, Cumulative share of a genus’ culprit specimens against species rank within that genus (log_10_ x axis), eight genera, counted per specimen. Filled circle, top-ranked species; legend entries give the genus, its species detected anywhere in the corpus and the share carried by its leading species. b, Culprit rate per detected specimen for a genus’ leading species (vermilion) against its remaining species pooled (grey), log axis; the printed multiple is their ratio. c, Within-unit AUC separating culprit from non-culprit detections of the same species at the same site. Row, one sequencing index; point, one species × site unit; outline, kernel density; tick, median. Blue above the row, the 168 units of the ≤2024 cohort; vermilion below, the 61 recurring in 2025 Q1; dashed line at 0.5, no separation. Right-hand column, median and 95% confidence interval (Methods). d, Average precision over the same units, for each index alone and for four combined models. Square, single index; circle, combined model; heavy bar, 95% confidence interval; pale stem, the gain over that cohort’s causative prevalence (dashed and dotted lines). e, Clinician against model culprit rates per detected specimen, 35 species of eight genera in the 2025 Q1 cohort. Both axes share one inverse-hyperbolic-sine transform, so the dashed line is the exact identity. Marker area, specimens detected; vermilion, shell species; blue, core species. The model is fitted on the clinician labels, so a divergence describes the fit rather than testing the label.

Species identity and specimen source each contributed additional information. Within several clinically important genera, culprit calls were concentrated in a leading species^24–26^. *Pseudomonas aeruginosa* accounted for 91% of culprit specimens in its genus despite 187 detected species, with a culprit rate 284-fold higher than that of its pooled congeners (Fig. 2a,b). For the same species, rates also differed across sites. Among specimens in which *E. coli* was detected, it was judged causative in 37.4% of plasma, 11.8% of BALF and 3.1% of CSF specimens (q = 5.6 × 10⁻¹¹; Fig. 3a). *Stenotrophomonas maltophilia* was judged causative in 21.4% of BALF specimens in which it was detected, compared with 1.2% of CSF specimens. Across the testable species, 48 of 234 showed site heterogeneity at a false discovery rate below 0.05. These are associations with recorded clinical judgements, not estimates of intrinsic pathogenicity.

**Figure 3.**
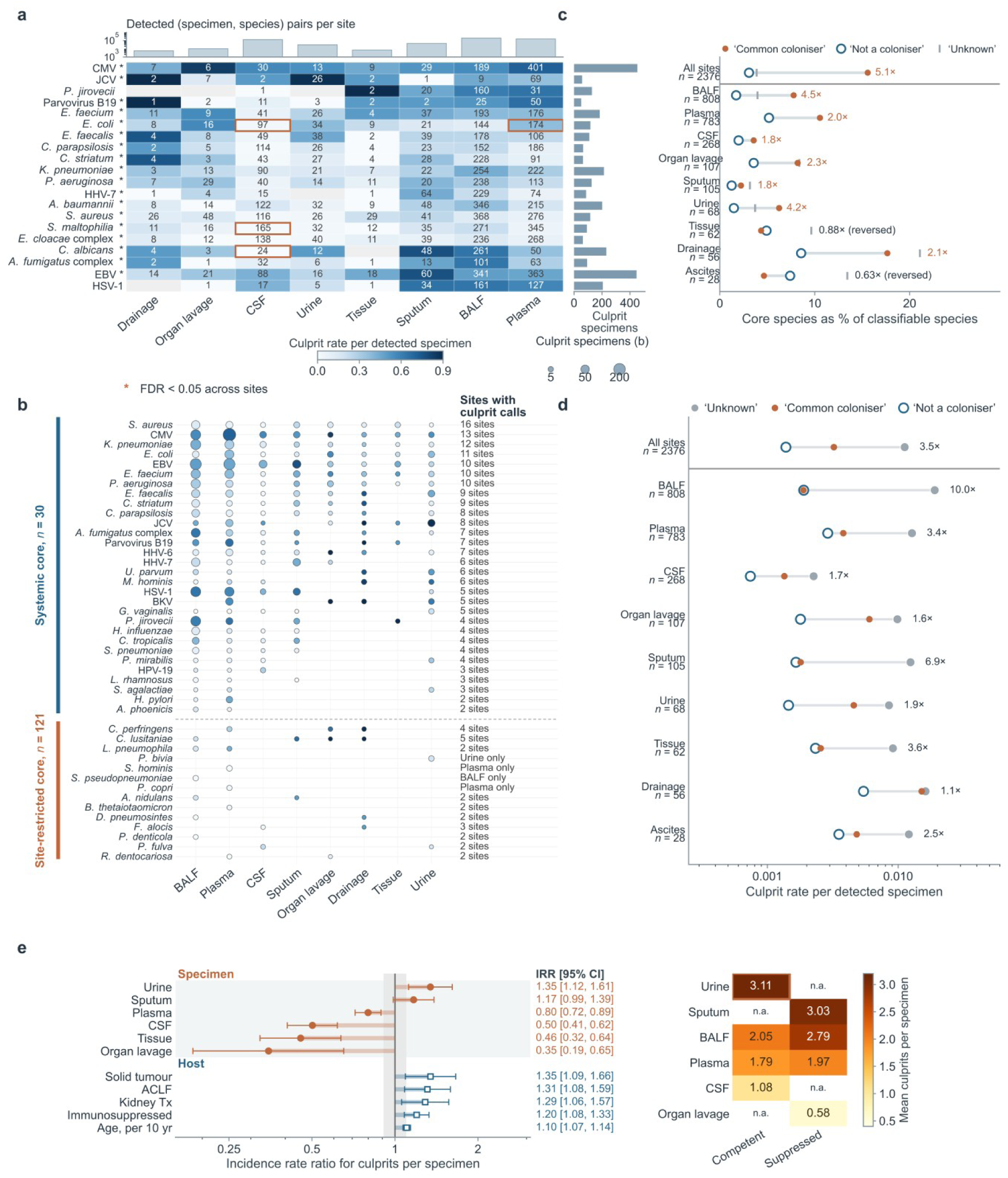
Specimen source and coloniser annotations in clinical adjudication. a, Culprit rate for the 20 species with the most causative specimens across the eight major body sites. Cell colour, causative specimens as a fraction of those in which the species was detected at that site (scale shared with b); numeral, the detected specimens behind that rate; grey, not detected. Marginal bars, detected (specimen, species) pairs per site and causative specimens per species. Asterisk, rate heterogeneous across sites (χ², Benjamini–Hochberg FDR < 0.05); vermilion outlines, the contrasts quoted in the text. b, The same calls split by the number of sites at which a species is ever causative: systemic (blue bracket, n = 30) against site-restricted (vermilion, n = 121). Dot area, causative specimens for that pair; dot colour, its rate on the scale of a. c, Curated coloniser label against the data-driven core/shell partition, computed within site (Methods). Filled vermilion, ‘common coloniser’; open blue ring, ‘not a coloniser’; grey tick, ‘unknown’; text, the ratio of the two labelled shares, Fisher exact test. d, Causative rate per detected specimen by knowledge-base label, on the strata of c; log scale. e, Determinants of the number of causative pathogens per specimen, as incidence rate ratios from one multivariable Poisson model (Methods). Filled circle, specimen-type term; open square, host term; bar, 95% confidence interval. Right, mean culprits per specimen by site and host immune status.

The curated coloniser annotation cannot be used on its own to judge that an organism is non-causative (Fig. 3c,d). In the pooled comparison, organisms annotated as common colonisers had a higher recorded culprit rate than those explicitly annotated as non-colonisers; within BALF, this distinction was less clear. An annotation describing common residence at a body site does not capture all opportunities for infection, and resident organisms can also be opportunistic pathogens^27–30^. Species never labelled causative in this dataset should likewise not be assumed harmless, because the labels cannot distinguish true absence of pathogenic relevance from overlooked calls.

Input-masking experiments provided complementary evidence about the model (Supplementary Fig. 2). Masking quantitative indices reduced average precision from 0.825 to 0.296 and increased false positives from approximately 2.0 to 7.44 per specimen. Masking organism identity reduced culprit recall, whereas masking the report annotation increased false positives to 3.39 per specimen with little change in recall. Masking the available clinical fields or sampling-site label had smaller effects. These experiments measure the model’s sensitivity to the supplied fields. As inputs may be redundant and clinical recording often incomplete, a small effect should not be interpreted as evidence that the corresponding clinical information is unimportant.

### Patterns of joint pathogen adjudication form species-level modules that vary across specimen types

Culprit counts were overdispersed and were better fit by a negative binomial than a Poisson model (Supplementary Fig. 3). In the earlier held-out cohort, sampling site was more strongly associated with culprit count than the recorded host factors, although the available clinical variables were incomplete (Fig. 3e). Mixing of bacterial, fungal, and viral culprits did not exceed the level expected under a permutation model that preserved class frequencies and each specimen’s culprit count (Fig. 4a,b).

**Figure 4.**
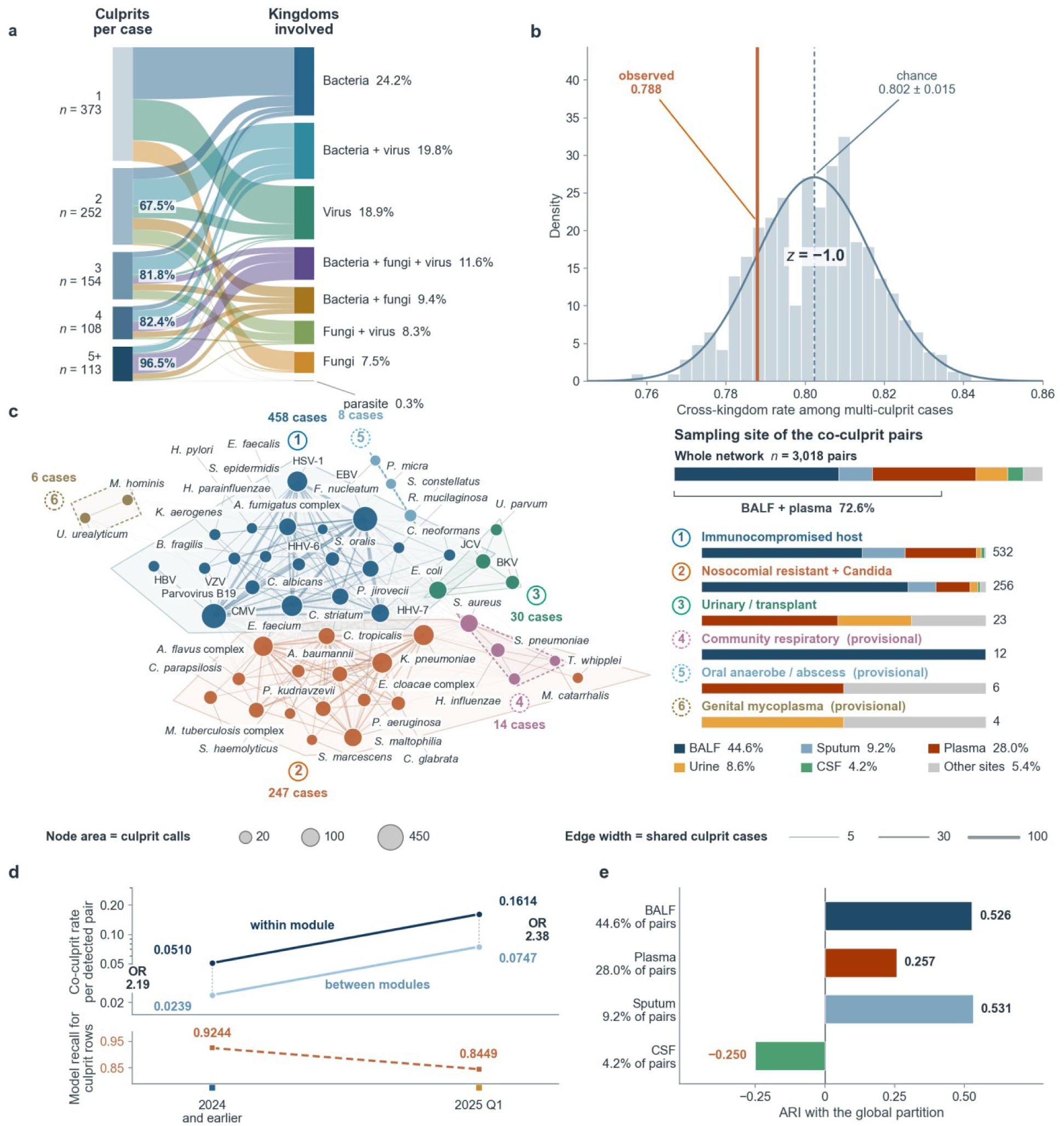
Species level patterns of coadjudication across specimen types. **a**, Organism-class composition of culprit-bearing cases against the number of culprits, ≤2024 cohort (n = 1,214 specimens; one case per specimen throughout). Ribbon width, cases; left nodes, culprit count; right nodes, the set of classes among a case’s culprits; blue figures, each group’s cross-class share. **b**, Label-reshuffling test of whether cross-class mixing exceeds chance, over the multi-culprit cases of the same cohort. Histogram, null from 1,000 reshuffles; solid curve, fitted normal density; dashed line, null mean; vermilion line, the observed value. The null holds the class composition of the culprit pool and each case’s culprit count fixed (Methods). **c**, Co-culprit network over the same cohort. Node, one pathogen; node area, causative calls; node colour, module (Methods); edge width, cases in which both were causative. Layout is force-directed and carries no quantitative meaning. Key, the sampling-site provenance of each module’s co-culprit pairs, with its within-module pair count at the right. Dashed outlines mark modules 4–6 as provisional (14, 8 and 6 clustered cases). **d**, Left axis, co-culprit rate per detected pathogen pair for same-module pairs (dark) against cross-module pairs (light); the connector label gives the odds ratio. Right axis (vermilion, dashed), model recall over causative rows in the same cohorts. **e**, Adjusted Rand index between the partition obtained inside one sampling site and the global partition of c, for the four sites large enough to support a site-specific network; each bar is labelled with that site’s share of all co-culprit pairs.

At species level, the co-adjudication network contained six candidate modules (Fig. 4c). The largest module grouped opportunistic fungi and herpesviruses, including cytomegalovirus, Epstein–Barr virus, *Pneumocystis jirovecii* and the *Aspergillus fumigatus* complex, with 458 co-occurring cases^31–36^. A second grouped organisms associated with hospital-acquired infections and *Candida*, with 247 cases, while a third included urinary and transplant-associated organisms, including JC and BK polyomaviruses, with 30 cases^37–40^. The community-respiratory, oral-anaerobe and genital-mycoplasma groupings were supported by 14, 8 and 6 cases and remain provisional^41–45^.

Although some organisms consistently co-clustered, the exact module partition was unstable. Across 500 specimen-level bootstrap samples, the median adjusted Rand index was 0.361 and six clusters recurred in approximately 39% of samples (Supplementary Fig. 4). Organisms within the original modules were more likely to be co-adjudicated in both the earlier and later cohorts, with odds ratios of 2.19 (95% confidence interval, 2.00–2.41) and 2.38 (2.17–2.61), respectively (Fig. 4d). The later enrichment supports the original grouping, but does not establish independent recovery of the same six modules.

The global network was dominated by BALF, plasma and sputum, which contributed 81.8% of co-culprit pairs. Site-specific partitions agreed more closely with the global partition in sputum and BALF than in plasma (Fig. 4e; Supplementary Fig. 5). The CSF graph contained only seven nodes and six edges across 268 specimens, which was insufficient to establish either a comparable module structure or a distinct infection ecology. These patterns warrant further clinical investigation, but should not be extrapolated across specimen types without additional support.

### Adjudication is partner-dependent, but supplying partner lists does not improve prediction

For selected organisms, the recorded culprit judgement was associated with the judgement of a specific co-detected organism. Among 105 evaluable within-module pairs, 49 showed positive associations at a false discovery rate below 0.10 (Fig. 5a). *S. maltophilia* was judged causative in 28.7% of specimens in which a co-detected *Acinetobacter baumannii* was causative, compared with 8.9% of specimens in which *A. baumannii* was non-causative (odds ratio, 4.1). Other examples included *A. baumannii* with *Klebsiella pneumoniae* (3.4), *Staphylococcus haemolyticus* with *Enterococcus faecium* (6.1), and *Candida parapsilosis* with *E. faecium* (4.9). Detection-opportunity-adjusted summaries showed that these relationships were asymmetric, but could not establish causal direction (Fig. 5b).

**Figure 5.**
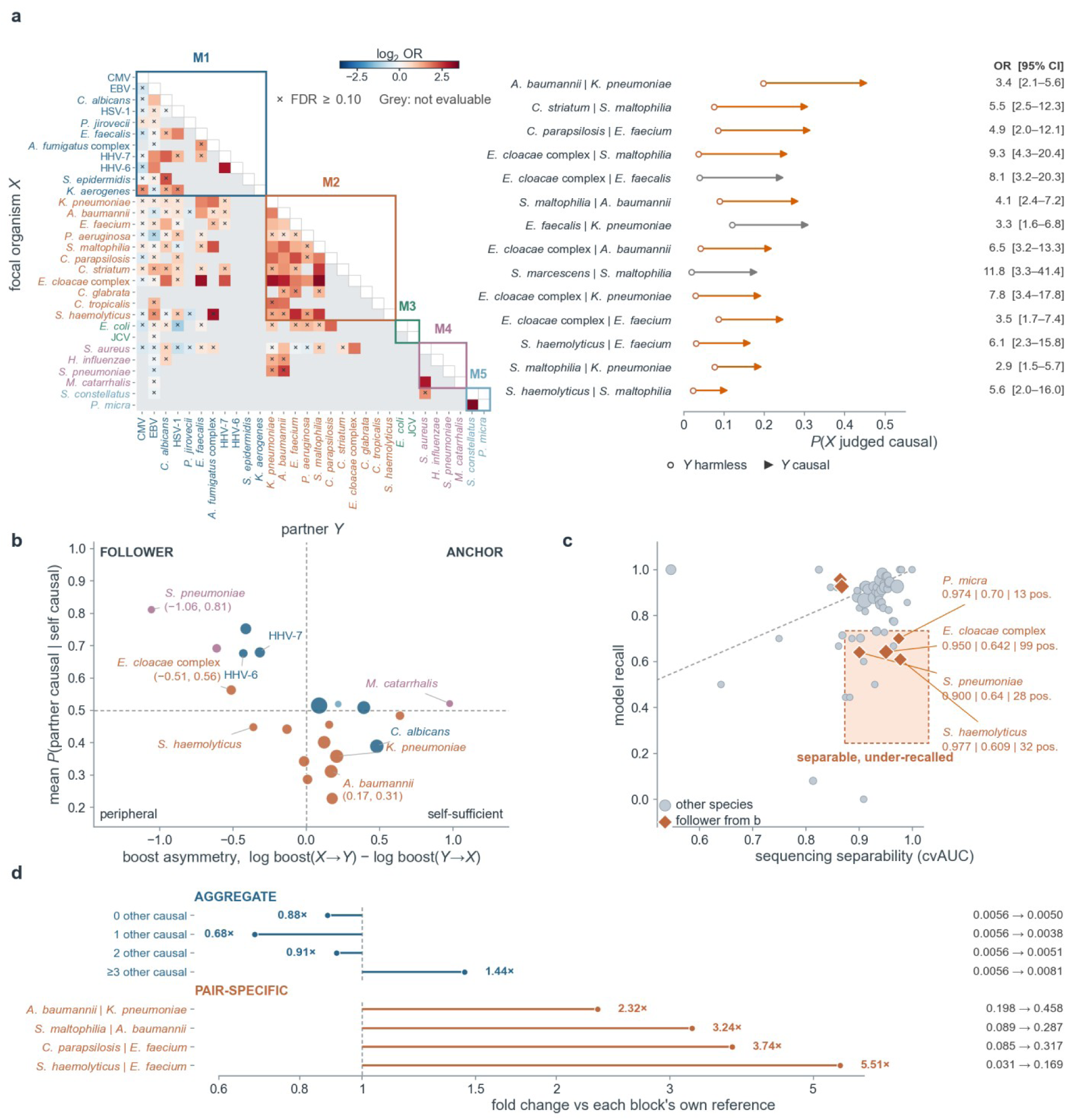
Partner associated adjudication and model recall. **a**, Left, pairwise conditional causative rates among the 30 organisms entering at least one evaluable co-detected pair. Cell, log₂ of the odds ratio for the focal organism X (rows) given the partner Y (columns), with co-detection held fixed (Methods); only the lower triangle is drawn, saturating at |log₂ OR| = 3.5. Crosses, evaluable but not significant (FDR ≥ 0.10); grey, too rarely co-detected; coloured boxes, the six modules of Fig. 4. Right, the significant pairs as absolute probabilities: open circle P(X a culprit | Y harmless), arrowhead P(X a culprit | Y a culprit), so arrow length is the absolute rise; grey arrows are cross-module. Right-hand column, the odds ratio with its 95% confidence interval. **b**, Leader–follower quadrant. x, detection-opportunity-corrected boost asymmetry averaged over an organism’s partners, positive values identifying anchors (Methods); y, the mean probability that a partner is a culprit given that the organism itself is. Point area, culprit calls; colour, module as in a. **c**, Point, one of 61 evaluated entries with both a separability estimate and a model recall; point area, culprit calls. x, cross-validated AUC for separating culprit from non-culprit detections of that species from sequencing metrics alone; y, the model’s recall at the fixed decision rule. The dashed line marks equal numerical value only: AUC is threshold-free and recall is not, so it is a reading aid, not a reference expectation. **d**, Aggregate (blue) and pair-specific (red) effects on one logarithmic axis of fold change relative to each block’s own reference. The blocks use different denominators; their relative fold changes do not isolate the contribution of partner identity from other factors (Methods).

Some organisms showing partner associations, including the Enterobacter cloacae complex, S. haemolyticus, Parvimonas micra, and Streptococcus pneumoniae, were also incompletely recovered by the model despite high separability from quantitative indices (Fig. 5c). Their cross-validated AUCs ranged from 0.900 to 0.977, while model recall ranged from 0.61 to 0.70. These measures capture different properties, so they do not establish that partner information is necessary for prediction. Aggregate associations with the number of other culprits were non-monotonic in the earlier cohort, and candidate-panel size also affected the proportion of records labelled causative (Fig. 5d; Supplementary Fig. 6). Because these summaries use different denominators, they cannot separate the effect of partner identity from that of overall burden, sampling site, or host background.

We tested two ways of adding information from other organisms in the specimen. A list of 12 co-detected organisms did not improve performance and captured the reference co-culprits in only 41.9% of cases. A two-stage approach supplied first-pass culprit predictions as context for a second assessment. In 2025 Q1, culprit recall fell from 84.49% to 75.96%, and recovery of every reference culprit in a specimen fell from 67.89% to 51.04% (Supplementary Fig. 7). The largest decline occurred in the reference-label stratum with no other culprit, from 88.5% to 46.0%, although this stratum does not necessarily correspond to an empty predicted context. Five of six selected organisms also had lower recall. These implementations therefore failed to convert the observed associations into better predictions.

### Model evaluation against blinded clinical re-adjudication and microbiological evidence

Because the reference labels are expert judgements rather than a microbiological ground truth, their reproducibility must be measured directly. The 215 mNGS reports from March–April 2025 within the 2025 Q1 cohort served as part of the cross-time evaluation and were additionally re-adjudicated by another independent clinical team, blinded to the original culprit-pathogen calls (Fig. 1d), providing a measurable bound on label reproducibility. The original and repeat assessments identified exactly the same set of culprit pathogens in 77.2% of specimens, with Dice/F1 agreement of 0.898 and Jaccard similarity of 0.815. Disagreements were concentrated in particular organisms, including Epstein–Barr virus and *Escherichia coli*.

We compared model predictions with both clinical assessments (Fig. 6a,b). Among the 359 candidate records judged causative by both teams, model recall was 90.5%, compared with 71.1% and 70.6% for calls made by only one of the two readers. However, the model classified 589 records as causative, compared with 397 and 376 for the readers, and 225 model-positive records were judged non-causative by both. Model–reader Dice agreement was 0.714 and 0.698, below the inter-reader value of 0.929 on these candidate records. The latter differs from the pathogen-name agreement reported above because calls without a corresponding candidate record are excluded.The model thus recovered most shared expert calls but also generated many additional positive predictions.

**Figure 6.**
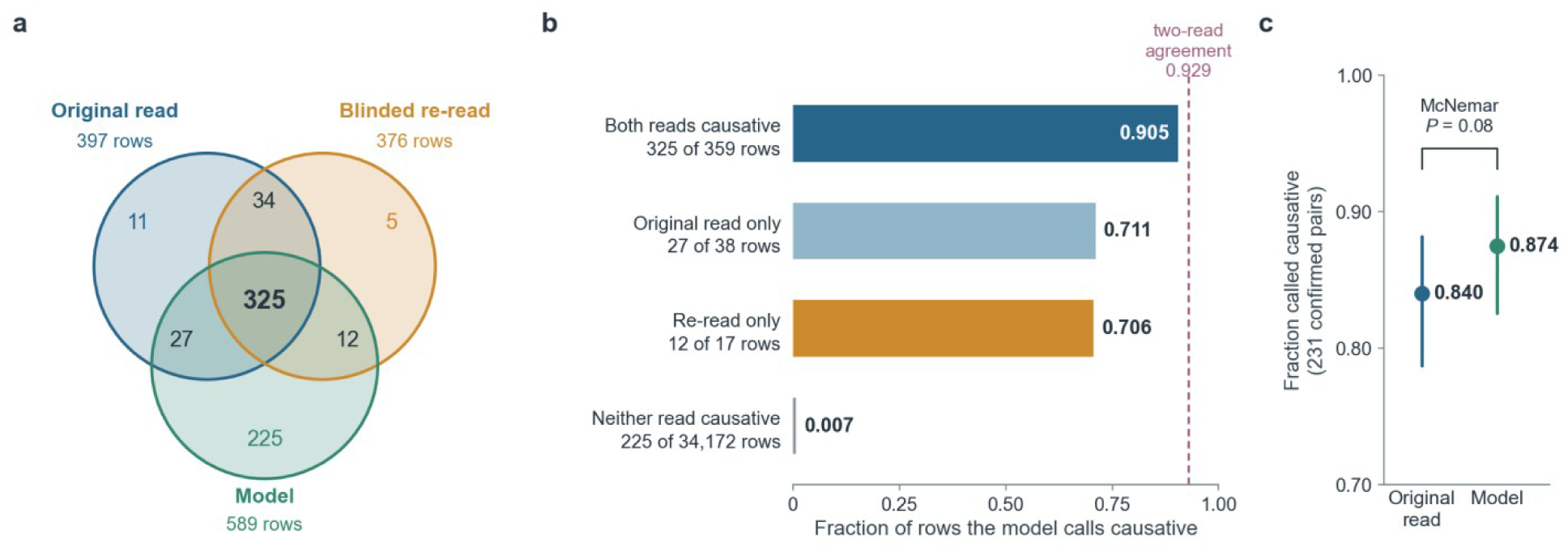
Comparison of model predictions with independent expert review and microbiological evidence. **a**, Three-set overlap between the causative calls of the original read, of a blinded re-read by the hospital team, and of the model at the fixed decision rule, over every candidate detection row of the 215 re-adjudicated cases (34,586 rows). Blue, original read (397 rows); orange, blinded re-read (376 rows); green, model (589 rows); 33,947 rows are causative in none of the three. Calls that could not be placed on any candidate row, almost all of them RNA pathogens absent from this DNA-only cohort, are outside this panel (Methods). **b**, Model recall within each stratum of reader agreement, on the same rows; the bottom bar is a false-positive rate rather than a recall. Dashed line, inter-reader Dice agreement at the candidate-row level. This statistic and recall measure different quantities and should not be interpreted as a model performance ceiling. **c**, The same comparison against an independent microbiological reference: organisms confirmed on the same specimen by culture, smear or nucleic-acid testing (n = 231 organism–specimen pairs). Error bars, Wilson 95% confidence intervals; the paired comparison is an exact McNemar test.

An additional comparison used 231 organism–specimen pairs with same-specimen confirmation by culture, smear or nucleic-acid testing (Fig. 6c). The original reader judged 194 of these organisms causative (84.0%; 95% confidence interval, 78.7–88.2%), and the model judged 202 causative (87.4%; 82.6–91.1%; exact McNemar P = 0.077). These tests corroborate organism presence rather than causation, and this comparison does not establish equivalence between the model and clinical readers. Together, these evaluations show that the model’s value lies not in replacing expert adjudication, but in being highly reliable where experts agree and concentrating its disagreements where experts themselves cannot agree—precisely the region where clinical review most needs support.

## Discussion

This study examines the interpretation step that connects a clinical mNGS report to a judgement about infection. By linking complete candidate panels to recorded culprit calls, we established a reference for evaluating both the information associated with adjudication and the behaviour of an automated system. The language model recovered many calls shared by independent expert readers, but produced substantially more positive calls. Quantitative evidence was most informative when considered alongside organism identity and specimen source, while patterns of co-adjudication revealed additional complexity in reports with multiple candidate pathogens. Together, these findings show that candidate-review assistance warrants evaluation, and that recovery of reference calls alone does not establish clinical usefulness.

The clinical question is whether a tool helps the reviewer identify relevant organisms without encouraging overinterpretation. The model’s additional recovery relative to a tree model was accompanied by additional false positives, and the expert comparison showed that excess calls remained a substantial source of disagreement. These results support investigating a workflow in which predictions are reviewed alongside the underlying report and clinical information. They do not establish which model or operating point offers the best clinical balance. That balance depends on the consequences of an overlooked organism, the burden of checking additional candidates and the evidence available to resolve uncertainty. High specificity across many non-causative candidates can still be accompanied by several additional positive calls in an individual report, so specimen-level summaries are important for clinical interpretation.

The findings also support organism- and specimen-specific interpretation of sequencing evidence. A high read count or coverage value has a different meaning when the organism and sampling compartment change, and an organism’s inclusion in a coloniser list does not determine its relevance to the current episode. The substantial differences between congeners and between sites suggest that broad organism categories can obscure useful information. At the same time, the labels record the decisions of clinicians who had access to these measurements. The associations therefore describe clinical adjudication and should not be treated as independently established diagnostic thresholds. CSF illustrates the need for separate assessment: its low culprit prevalence, lower model recall in the earlier cohort, and sparse co-adjudication network make extrapolation from respiratory-dominated results difficult.

The co-adjudication analyses extend this interpretation to reports containing several possible pathogens. Certain organisms were judged causative together more often than detection alone would predict, and some associations were observed again in the later cohort. These findings may help formulate hypotheses about which combinations deserve closer study. However, shared sampling site, host susceptibility and clinical reading practices can generate such patterns. Co-adjudication does not establish simultaneous infection or biological synergy, and the relationships described here cannot directly guide combination treatment^46,47^. The failed context experiments make a practical point: additional information must be evaluated in the form in which it would actually be supplied and used. The present results do not show that naming other predicted culprits improves the decision, nor do they identify a unique mechanism for the deterioration.

Several limitations affect interpretation. Reference labels reflect treating-team judgement, and a same-hospital blinded review cannot exclude shared clinical biases. Clinical context was incompletely recorded, and the later cohort differed in library preparation and label construction, so its performance decline cannot be attributed to time alone. Repeat admissions and tests represent legitimate clinical events, but observations from the same patient may be correlated. The available patient-overlap sensitivity analyses showed small recall changes in the evaluated subsets; the earlier analysis covered 607 culprit-bearing specimens and does not establish the effect across the complete held-out cohort. The study was conducted at one centre, module membership was stable only for a core of organisms, and small modules require further evidence. Finally, we evaluated agreement with reference judgements rather than the effect of model assistance on clinicians, antimicrobial decisions or patient outcomes.

The clinical challenge in mNGS is deciding which detected organisms explain the patient’s current infection. The cohort and benchmark presented here serve that question: by linking complete detection panels to expert culprit determinations, they make a previously unstudiable decision step evaluable and provide a reusable cohort for further model development on this task. Our results also indicate that similar studies should further test whether AI-assisted models can improve the consistency and timeliness of adjudication without increasing unnecessary positive interpretations, and ultimately examine their effect on clinical decisions. These are the outcomes that will determine whether automated support genuinely helps clinicians.

## Methods

### Cohort and data sources

The corpus comprises mNGS panels from 4,855 specimens tested at a single tertiary centre in two cohorts (2024 and earlier: 3,693 specimens, of which 2,479 development and 1,214 held-out test; 2025 Q1: 1,162 specimens), covering 19 sampling sites and 1,333,867 adjudicated organism-level rows (719,426, 431,566 and 182,875 in the three splits). Specimens are the unit of analysis and every count is a specimen count unless stated otherwise; one specimen is one case throughout, and one patient may contribute several specimens. Patient identity was reconstructed inside the hospital from admission number and name, giving 889 patients for the 1,162 specimens of 2025 Q1 and 2,607 patients for the 3,405 specimens of the pooled ≤2024 cohort that enter the within-unit analysis, obtained by resolving the inpatient-number and name fields jointly (860 distinct inpatient numbers, 57 specimens with no number recorded, 26 patients readmitted under two numbers); patients sharing a name would be merged by this rule, so the figure is a lower bound. The study size is reported as specimens rather than distinct patients.

Sequencing used the laboratory’s clinical DNA and RNA metagenomic assay, which the consent document specifies as covering more than 17,500 microbial species and their resistance and virulence genes at a stated limit of detection of approximately 100 to 1,000 copies per mL. Both cohorts are reported against the same reference database (≈1.7 × 10⁴ species), so no comparison in this paper spans a change of reference database. Median candidate panel size is 186 rows per specimen in the ≤2024 development split, 243.5 in the ≤2024 held-out split and 102 in 2025 Q1.

The development split was used for model fitting and also entered the within-unit analysis in Fig. 2c, d. Other descriptive analyses used the held-out cohort, as specified below. Within-unit discriminability (Fig. 2c, d) is computed on the pooled ≤2024 cohort, since it requires (species, site) units with at least five causative and five non-causative detections and the split alone does not supply enough of them. Analyses of co-adjudication structure and culprit multiplicity used the ≤2024 held-out split (1,214 specimens), alone or pooled with 2025 Q1. Model evaluation used the held-out and later cohorts. The ≤2024 development/held-out split was made by specimen rather than by patient, so the two splits are not patient-disjoint. The overlap is measured rather than assumed, and is reported below with a sensitivity analysis; the 2025 Q1 cohort, drawn from a later period, is the generalisation test.

Per specimen, sequencing output was taken from per-class detection tables (bacteria, fungi, viruses, parasites) and clinical context from the structured clinical record together with parsed free-text run-day notes. Detection tables are GB18030-encoded; 34% of a 400-file audit failed to decode under GBK, and decoding with error replacement silently corrupts organism names, so GB18030 decoding is mandatory. Reported fields per row are smrn, species and genus relative abundance, CovRate, within-genus species rank, genus detection rank, Gram status, pathogen description and coloniser annotation. One reporting convention requires correction: the field labelled “species detection rank within the sample” is in fact the rank within the genus, capped at 10. Genus detection rank was reconstructed as a dense rank over the pooled bacterial, fungal and parasite tables by maximum within-genus smrn, with viruses ranked in a separate pool; this matches the reported field in 96.5% of rows against 59.8% when each class is ranked separately. Viral rows lack a genus field and are assigned genus = species with genus-level quantities set equal to species-level quantities. Patient background, where recorded, comprises age, sex, immune status, comorbidity group and admitting diagnosis; recording is incomplete and reaches 100% missingness for age and sex in 2025 Q1.

### Ethics

This study analysed only the structured report files generated by clinical metagenomic testing that had already been performed as part of routine patient care. No specimens were collected, handled or consumed for the purposes of this work, no additional sampling was carried out, no raw sequence data were accessed, and no intervention of any kind was applied to any patient. The analysis dataset consists of per-organism reporting fields, the coarse clinical descriptors listed in Methods and the treating team’s recorded causative-pathogen calls, and contains no directly identifying information. Where an analysis required rows to be grouped by patient rather than by specimen, the grouping was derived inside the hospital from the clinical records by clinical staff, and only the resulting anonymous grouping key was released into the analysis dataset.

The institutional review body of Ren Ji Hospital, Shanghai Jiao Tong University School of Medicine determined that ethics approval and filing were not required for the analyses described here. Clinical metagenomic testing was ordered and performed as a diagnostic service under the hospital’s standard consent process for high-throughput pathogen sequencing.

### Label definition and validation

The prediction target is row-level causative status: whether the treating team named this organism in the causative-pathogen fields of the clinical record for this specimen. Causative-pathogen strings were NFKC-normalised, split on Chinese and Latin punctuation, stripped of trailing read counts and matched against the Chinese name, subtype, type and species fields of the detection table, with a unique-substring fallback. In 2025 Q1, 2,164 of 2,350 recorded causative entries (92.1%) matched a detection row; the 186 unmatched entries are almost all RNA pathogens, which have no detection row because that cohort contains DNA libraries only, together with eight M. tuberculosis complex entries recorded at complex rather than species level.

Because the earlier cohort’s labels derive from an independent database field, the derivation was validated by applying it in reverse to that cohort (1,214 samples, 431,566 rows): precision 0.9731, recall 0.8594, and an overall culprit rate 0.88× that of the original field. The derivation missed some reference culprit labels; consequently, some apparent model false positives may represent unlabelled culprit calls. This comparison does not provide an independent biological truth standard. Before merging, culprit rows are 4,625 of 719,426 candidate rows in the ≤2024 development split (0.643%), 2,738 of 431,566 in the ≤2024 held-out cohort (0.634%) and 2,379 of 182,875 in 2025 Q1 (1.301%). Across the 2,376 adjudicated specimens of the two analysed cohorts the mean is 1.93 culprit rows per specimen, 83% of specimens carry 0–3 and the maximum observed is 18.

Organisms are named by the Latin binomial carried by the reference database rather than by the Chinese report name. The mapping is not one-to-one: several database species may share one Chinese name at genus level, in which case the genus is quoted and the species left unspecified (for example one Chinese name shared by six Melampsora entries, reported as Melampsora sp.). The coloniser annotation is site-conditional rather than a lookup: at respiratory sites “respiratory coloniser” maps to yes and “non-respiratory coloniser” to no; elsewhere “background organism” maps to yes and “non-commensal” or “respiratory coloniser” to no; unresolved entries are unknown.

### Inter-rater reliability of the label

215 cases of March–April 2025 were re-adjudicated by another independent hospital team, blinded to the original causative-pathogen calls and working from the same reports. Agreement is reported at three levels, all with culprit names split and normalised by the rule the evaluation-set builder applies. At case level, whether the case carries any culprit at all: 157 cases positive in both reads, 55 negative in both, 2 positive only in the original read and 1 only in the re-read, giving agreement 0.986 and Cohen’s κ = 0.964; that κ rests on 3 discordant cases and is quoted as a secondary statistic. At pathogen level, treating each read as a set of (case, pathogen) culprit calls: 360 shared calls, 50 unique to the original read and 32 unique to the re-read, giving Dice/F1 0.898, Jaccard 0.815 and exact set agreement in 77.2% of cases, with a mean of 1.91 culprits per case in the original read against 1.82 in the re-read. At the level of candidate detection rows, which is what the benchmark scores (reported with Fig. 6): 359 rows called by both reads, 38 by the original read only and 17 by the re-read only, giving a row-level Dice of 0.929, while 40 original-read and 41 re-read calls cannot be placed on any candidate row and are excluded from that level. Per-pathogen disagreement rate is the fraction of cases containing that pathogen in which the two reads differ on its culprit status, reported for every pathogen called in at least 11 of the 215 cases (Fig. 1d), and the per-call table is released as source data. The same 215 cases were used to test whether the model tracks the two readers’ consensus or the original reader’s idiosyncratic calls: every candidate row was scored at the fixed decision rule and partitioned into rows called causative by both readers (359 rows, recall 0.905), by the original reader only (38, 0.711), by the re-reader only (17, 0.706) and by neither (34,172, 0.007). The model saw only the original reader’s labels in training.

### Counting unit

All species-level frequencies are counts of specimens, not of report rows, and are computed within sampling site. The taxonomic normalisation step below collapses closely related database entries onto one reporting unit, so when two or more constituent entries are reported from the same specimen the collapsed unit leaves several rows for what is biologically one detection, each of which clinicians adjudicate; row-level tabulation therefore double-counts these organisms. Across the 2,376-specimen pooled cohort the effect on detections is negligible (614,441 rows against 611,409 distinct specimen–species pairs, +0.5%) but the effect on culprit calls is not (5,117 rows against 4,597 culprit pairs, +11.3%), because merged entries are enriched among adjudicated pathogens. The inflation is confined to 7 of the 4,293 species detected in ≥20 specimens, with row-to-specimen ratios M. tuberculosis complex 5.24×, torque teno virus 2.14×, *E. cloacae* complex 1.95×, EBV 1.47×, M. avium complex 1.43×, A. flavus complex 1.33× and *A. fumigatus* complex 1.08×; every other species maps 1:1. De-duplicating to the specimen level changes 441 of the 11,243 (species, site) cells behind Fig. 3a (3.9%) and moves rates in either direction, depending on whether the duplicated rows were adjudicated as causative: the culprit rate of EBV in BALF falls from 0.627 to 0.487, whereas that of the M. tuberculosis complex in BALF rises from 0.469 to 0.632, taking the latter out of the twenty species with the most culprit calls in favour of parvovirus B19. Aggregate conclusions are unaffected (cross-site heterogeneity 48/234 against 49/235 at row level; systemic to site-restricted ratio 30:121 either way), but any per-species figure quoted for one of those seven organisms must be taken from the specimen-level tabulation. Pooling sites would additionally contradict this paper’s own conclusion that causative status is a (species × site) property, and a row-level pooled tabulation lets specimens with large panels dominate every statistic, since the rows of one specimen are not independent observations. Panels drawing on patient background (Fig. 3e) are computed on the ≤2024 cohort alone (1,214 specimens), the only period with near-complete background fields.

### Repeated specimens and patient overlap

Patient identity was reconstructed as the connected component of admission number and name: an admission number can be missing and a patient can be issued a new one on re-admission, so neither field alone is a valid key. The derivation was performed inside the hospital and only the resulting anonymous grouping key entered the analysis dataset. Every specimen of all three splits resolved to a patient. The ≤2024 development split holds 2,479 specimens from 1,940 patients, the ≤2024 held-out split 1,214 specimens from 972 patients, and 2025 Q1 1,162 specimens from 889 patients, that is 1.25 to 1.31 specimens per patient. Because the ≤2024 split was made by specimen, 125 patients contribute to both sides of it, which is 12.9% of held-out patients and accounts for 159 of the 1,214 held-out specimens (13.1%). The later cohort is not fully disjoint either: 42 patients of 2025 Q1 (4.7%) also appear in the ≤2024 corpus, accounting for 52 specimens (4.5%).

The consequence for the reported metrics was measured on the specimens carrying per-case model output, which is the whole 2025 Q1 cohort (1,162 specimens) and the 607 culprit-bearing held-out specimens of the deep-panel analysis. Removing every specimen whose patient also contributed to the training split moves row-level culprit recall by +0.0061 in the held-out split, from 0.9116 to 0.9177, and by −0.0012 in 2025 Q1. Keeping only one specimen per patient moves it by −0.0006 and −0.0063 respectively. In the held-out split the shared-patient specimens are in fact harder, not easier: recall on them is 0.8706 against 0.9177 on the rest, indicating no recall advantage for overlapping patients within this evaluated subset. The earlier sensitivity analysis does not cover the complete held-out cohort. The script that performs this analysis is released with the benchmark.

### Taxonomic artefact merging

Species-level frequency and co-adjudication analyses used a fixed rule table to merge three classes of reporting artefact: multi-granularity reporting of one agent (for example EBV as untyped, type 1 and type 2, which unmerged show mutual lift 5.1–8.7); species complexes (M. tuberculosis complex including M. bovis, M. caprae, M. orygis and M. africanum, M. avium complex, A. flavus, *A. fumigatus* and *E. cloacae* complexes, whose unmerged M. tuberculosis members show lift 23,522 at Kirkwood K = 0.01); and renaming or splitting (*K. pneumoniae* / quasipneumoniae / variicola, which generate spurious mutual exclusivity). Merging reduced per-case culprit counts by 11.7% (≤2024) and 8.4% (2025 Q1), leaving 2,417 and 2,180 adjudicated culprit rows. The scale of the artefact was quantified before merging on the pooled 2,376 samples by lift, the observed frequency of two or three organisms being called culprits in the same sample divided by the frequency expected under independence (Supplementary Fig. 1): over the 544 pairs with at least 3 co-culprit events and at least 5 culprit calls per member, median lift is 2.28 for the 500 genuine biological pairs, 1.28 for the 3 rename/split candidates, 5.3–8.6 for the 3 multi-granularity pairs and 148 for the 38 species-complex pairs.

### Model fine-tuning and evaluation

The model is Qwen3-8B^23^, supervised fine-tuned on the row-level adjudication task over the ≤2024 development split (2,479 specimens, 719,426 adjudicated rows, of which 646,600 enter the fitting file) with hard-negative mixing and coloniser-aware prompting. The fitting set is not the natural row distribution: culprit rows are upsampled and retained negatives are drawn preferentially from hard candidates, so the training file contains 32,330 culprit rows, exactly 5.00% of its rows, against a natural prevalence of 0.634% in the held-out split, an enrichment of about 7.9×. The hard-negative mixture scores every causative row on rule disagreement, coloniser-annotation conflict, low-signal positivity, diagnosis match and species-level label mixing and duplicates it 1 + min(score, 6) times, and scores every harmless row on rule over-call, high-signal negativity and label mixing, retaining it by weighted sampling without replacement with weight 1 + 2 × score. The 0.30 causative fraction is the target of that sampling stage; the retained harmless rows are then returned to the fitting file, whose final composition is the 32,330 culprit rows among 646,600 quoted above, that is 5.00%.

Fine-tuning used low-rank adaptation (rank 8, α = 32, dropout 0.05, no bias term, on the query, key, value, output, gate, up and down projections; 21.82 M trainable parameters, 0.266% of the 8,212.6 M of the base model), AdamW at learning rate 5 × 10⁻⁵ with a cosine schedule and 0.03 warm-up ratio, one epoch of 5,052 steps, per-device batch 4 with gradient accumulation 4 over 8 Ascend 910 accelerators (effective batch 128), maximum sequence length 1,024 tokens, bf16 precision, gradient checkpointing and seed 42 for both model and data order. The loss is a token-level focal loss with class-specific α on the two answer tokens; γ and α were supplied through the launcher environment and are not serialised in the run snapshot. The actual focal-loss γ and class-weight α values were not preserved in the available run snapshot and require verification against the training records. The checkpoint at step 5,052 was used for every evaluation reported here and the available configuration is to be included with the code release.

Prediction uses fixed argmax decoding, the decision rule being harmful logit > harmless logit. Metrics are defined at two levels: row-level culprit recall is tp/(tp + fn) over all candidate rows and row-level harmless-row recall is tn/(tn + fp); false positives per case is the total number of false-positive rows divided by the number of evaluated cases; case-level complete recall is the fraction of culprit-bearing cases in which every culprit row of that case is recovered (1,000 of 1,214 cases in the ≤2024 held-out split and 819 of 1,162 in 2025 Q1 are culprit-bearing). Headline metrics are, for the ≤2024 held-out split, culprit recall 0.9244, harmless-row recall 0.9943, case-level complete recall 0.8400 and 1.998 false positives per case, and for 2025 Q1, 0.8449, 0.9937, 0.6789 and 0.980. Recall is additionally reported by sampling site with 95% Wilson intervals, running in the held-out split from 0.960 in BALF (1,131/1,178 culprit rows), 0.969 in sputum and 0.930 in plasma down to 0.591 in CSF (104/176), and in 2025 Q1 from 0.931 in sputum and 0.890 in BALF down to 0.806 in plasma, 0.778 in organ lavage and 0.718 in CSF. Parasite rows are reported as a separate stratum and excluded from aggregate metrics, that stratum carrying only 4 culprit calls over 4 species. Cohort-wide metrics are reported as point estimates. They are exact for the cohorts as constituted, but any expectation about future patients carries sampling uncertainty that these figures do not express; the site-level Wilson intervals and the cluster bootstrap of Fig. 2 give the scale of that uncertainty where it was estimated.

### Reference baselines

Sixteen models were scored on the same candidate rows, the same labels and the same two cohorts as the fine-tuned model. They are: two null models (all rows harmless; a random draw at cohort prevalence); four single-index threshold rules, on species-specific reads, species relative abundance, genome coverage and within-genus species rank, each threshold fixed on the development split by maximising F1 there (smrn 355, species relative abundance 44.85, coverage 6.91; the rank rule admits no useful threshold); two historical-rate priors, the empirical culprit rate of the species and of the (species × site) pair, estimated on the development split; a logistic regression and a gradient-boosted tree ensemble over the nine quantitative indices with sampling site and specimen type; three further tree ensembles adding species identity and then the report text fields, with the diagnosis and organism-description fields dropped in turn; and the same full tree ensemble fitted on the causative-enriched file used for language-model fine-tuning. Average precision and AUROC summarise performance across thresholds. Precision, recall, F1, false positives per case and case-level complete recall were calculated at each model’s operating point on identical evaluation rows. These comparisons therefore reflect both model performance and operating-point choice. Text fields were additionally audited for label leakage, by measuring for each candidate leak channel how many rows it flags and what fraction of those are causative. The only channel whose positive predictive value rises materially above prevalence is a diagnosis field naming the same genus as the row: it flags 85 of 431,566 held-out rows (0.02%) at a positive predictive value of 0.13, so it can account for at most 11 of the 2,738 causative rows (0.4%). Every channel with appreciable coverage carries a positive predictive value at or below 0.017 in both cohorts, that is at or below three times prevalence. This audit did not identify a high-coverage direct label channel; it cannot exclude every form of information leakage.

### Statistics and reproducibility

Within-unit discriminability and threshold rules. The unit is a (species, body site) pair with at least five culprit and five non-culprit detections, counted per specimen. Within each unit every candidate index is scored by AUC and by average precision, which summarises the precision–recall trade-off across thresholds under class imbalance. Every interval in Fig. 2c, d is a 95% percentile interval from 1,000 cluster bootstrap replicates that resample patients, each drawn patient carrying all of its specimens. The core-and-shell statistic below instead resamples units, 2,000 times; the two are stated separately because the resampling unit differs. Combined rules are logistic models over the five reported indices, fitted and evaluated by 5-fold cross-validation stratified on the outcome with folds drawn over units, so that no unit contributes to both fitting and evaluation; the standardised variant replaces each index by its z-score inside its own unit, and the historical-rate variant adds the empirical culprit rate of that unit estimated on the fitting folds only. The same procedure is applied to the 61 units that recur in 2025 Q1.

Core and shell species. Core species are those with at least one clinician causative call; shell species are detected congeners with none. Two known biases bound every statement built on this partition: it is estimated on the same corpus in which the classes are then compared, and because a species enters the core as soon as it is called causative once, frequently detected species reach the core more readily than rare ones at an equal per-detection rate. The classes are therefore used descriptively and not as an independent variable in any inferential claim; the cleaner design would define the classes on the ≤2024 cohort and estimate causative rates on the later one. Where shell and core rows are compared directly, the unit is a (specimen, genus) pair with at least two shell rows and at least two non-culprit rows of core species, so that both sides are non-culprit rows and the sampling site is held fixed; each unit contributes one paired difference and the summary statistic is the fraction of units in which the difference is positive, with a 95% confidence interval from 2,000 bootstrap resamples of units, and is released as source data. Within-unit separation is also expressed as an AUC, a linear rescaling of Cliff’s δ^48^. Genus detection rank is excluded because it is constant within a unit.

Site heterogeneity and breadth. For each core species observed at two or more sites, culprit proportions were compared across sites by Fisher exact or chi-square test as appropriate, with Benjamini–Hochberg control of the false discovery rate^49^ across species. The testable set comprises global core species reaching n ≥ 10 detected specimens in at least two major body sites with at least one culprit call among them; 48 of 234 testable species (20.5%) are heterogeneous at FDR < 0.05 and 497 of 11,243 (species, site) cells are core (4.4%). Systemic core species (n = 30) have culprit calls in every body site in which they are sufficiently observed; site-restricted core species (n = 121) have culprit calls in exactly one such site.

Coloniser knowledge base. The curated field was cross-tabulated against the data-driven core / shell / unclassifiable partition, computed within each sampling site and per specimen: a species is core at a site if it is the culprit in at least one specimen from that site, shell if detected in at least 5 specimens from that site without ever being a culprit, and unclassifiable otherwise. Sites enter when both curated labels carry at least 25 classifiable species, which admits nine sites. Per-site shares, Fisher exact tests and culprit rates by label are plotted in Fig. 3c, d and released as source data.

Culprit count and its determinants. Poisson, negative binomial (NB2) and zero-inflated negative binomial models were fitted by maximum likelihood and compared by likelihood-ratio test (Poisson against NB2) and by the fitted mixing parameter (NB2 against ZINB). Variance explained by each categorical covariate was quantified as ε² from Kruskal–Wallis tests^50^. Multivariable Poisson regression of culprit count on sampling site, immune status, comorbidity and age group used BALF and immunocompetent status as reference, additionally adjusted for log candidate-panel size and clinical diagnosis group, with robust standard errors. Both the ε² decomposition and the regression were fitted on the ≤2024 cohort (n = 1,214), the only cohort with near-complete immune-status and comorbidity recording; the fitted incidence rate ratios and stratum means are drawn with their confidence intervals in Fig. 3e and released as source data.

Organism-class assortment. The observed cross-class fraction was compared with a permutation null in which organism labels are shuffled within culprit-count strata, preserving each case’s culprit number and the marginal organism frequencies, and reported as a z score against the permutation distribution (1,000 reshuffles). Permuting instead within each sample over that sample’s own detected candidates gives z ≈ +25; that null answers the different question of whether culprits are a random sample of detections and is strongly rejected because roughly 87% of detections are bacterial whereas viruses are enriched among culprits. The two nulls are not interchangeable and the result reported in the main text is the one that holds the marginal class composition fixed.

Co-infection modules. A co-culprit graph over species was built from within-case co-adjudication weighted by the raw number of shared culprit cases. A species enters the graph if it is adjudicated causative in at least 12 cases and a pair is joined if both are causative in at least 3 shared cases, giving 49 nodes. Communities were detected by resolution-parameterised modularity maximisation^51^ (Louvain local-moving^52^, up to 25 sweeps in randomised node order, best of 20 restarts by Q) at γ = 1 on the raw-count graph, which is the partition reported throughout. Alternative edge weights (Jaccard, φ, maximum conditional probability, log lift) recover essentially the same communities (ARI 0.773 for Jaccard and 0.734 for φ against the raw-count reference) at higher modularity (Q = 0.25–0.35 against 0.146), which is expected because normalised weights suppress the degree heterogeneity that raw counts inherit from species prevalence. The full sweep of five weightings × six resolutions (γ = 0.5, 0.8, 1.0, 1.3, 1.6, 2.0), together with spectral and average-linkage hierarchical clustering on the Jaccard distance at k = 3–10, is mapped in Supplementary Fig. 4c and released as source data. Stability was assessed by bootstrap resampling of cases with replacement (B = 500), rebuilding the graph on the resampled cases restricted to the original node set, refitting the partition with the same algorithm and weighting, and reporting the adjusted Rand index^53^ against the full-data partition together with per-node assignment stability Δ, defined as the difference between how often a species is co-assigned with its own reference module and how often with the others. Module–immune-status and module–comorbidity associations are reported as site-stratified Mantel–Haenszel odds ratios^54^, and the sampling-site provenance of each module’s co-culprit pairs is given in Fig. 4c.

Site-specific module structure. The same detection and comparison procedure was applied within each major sampling site (Supplementary Fig. 5). No single site carries enough specimens inside the ≤2024 cohort alone, so the site-specific graphs are built on the pooled two-cohort corpus (n = 2,376 specimens) and compared against the ≤2024 global partition of Fig. 4c; the site partitions are therefore estimated on more data than the partition they are scored against, so the comparison is descriptive and is not an independent external validation of the partition. The graphs are sputum (105 samples, 19 nodes, 72 edges), BALF (808, 46, 337), plasma (783, 37, 162) and CSF (268, 7, 6). Site-specific clusters supported by fewer than 4 clustered cases are omitted from the composition panel; the supporting counts are 53 and 11 for sputum, 185, 172, 42 and 20 for BALF, 219, 37, 13 and 10 for plasma, and 7, 7 and 4 for CSF, in plotted order.

Pairwise association and higher-order structure. All 890 evaluable species pairs were tested by one-sided Fisher exact tests in both directions (positive association and mutual exclusion) with Benjamini–Hochberg correction at FDR < 0.10. Triplet structure was assessed by the Kirkwood superposition approximation^55^, K being the ratio of the observed triplet frequency to that predicted from the three pairwise frequencies, so that K ≈ 1 indicates no irreducible three-way term. Module 1 was additionally tested against the alternative that it is two fused clusters by partitioning its members a priori into fungal and herpesviral sets and comparing within- and between-set co-culprit rates as an odds ratio with a z score.

Pair-conditional adjudication. Analyses were conditioned on co-detection: for each ordered pair (X, Y), only samples in which both organisms appeared in the panel were retained, and P(X causative | Y causative) was compared with P(X causative | Y detected but harmless), reported as an odds ratio with a z score and corrected across the 105 evaluable within-module pairs by Benjamini–Hochberg. Directionality was assessed with detection-opportunity-corrected gain ratios rather than by comparing P(X | Y) with P(Y | X) directly, since the latter ratio is identically the ratio of marginal prevalences. Where two absolute probabilities are shown, one direction must be chosen because the odds ratio is symmetric while the probabilities are not: the four pairs quoted in the main text are shown with the focal organism the member of lower baseline probability and every other pair in the direction with the larger relative rise, the reciprocal directions being released as source data. The four pair-specific estimates rest on 360 co-detected samples for *A. baumannii* | *K. pneumoniae*, 485 for *S. maltophilia* | *A. baumannii*, 159 for *C. parapsilosis* | *E. faecium* and 285 for *S. haemolyticus* | *E. faecium*. Single-row separability was evaluated for each (site, organism) unit with sufficient positives by 5-fold cross-validated AUC over sequencing indices only, and units with high cvAUC but low adjudication recall were flagged.

Aggregate-level control. The probability that a row is causative was tabulated against the number of other causative organisms in the same sample (0, 1, 2, ≥3) and expressed relative to the corpus baseline. The aggregate and pair-specific effects are conditioned on different things and their denominators are not comparable — the aggregate strata are per candidate row across the whole panel, with probabilities near 0.005, whereas the pair-specific estimates are per co-detected sample and restricted to organisms that are already frequent culprits, with probabilities of 0.03–0.50 — so the fold changes are presented descriptively on a common scale (Fig. 5d). Their relative magnitudes cannot isolate a partner-specific effect from site, host background or overall culprit burden. On the ≤2024 cohort the strata comprise 75,005, 131,575, 90,032 and 132,965 candidate rows and the fold changes are 0.88×, 0.68×, 0.91× and 1.44× of a baseline rate of 0.0056. The stratification is cohort-dependent: the gradient is non-monotonic in this cohort and monotonic in the pooled corpus (0.68×, 0.74×, 0.91×, 1.50×). The aggregate fold changes were ≤ 1.5, compared with pair-specific fold changes of 2.3–5.5 under their respective conditioning. These differently conditioned estimates do not exclude confounding of the pair-specific associations.

Panel-size power law. The per-row culprit prior was fitted as a power law in candidate count by ordinary least squares on log₁_0_ axes over the culprit-bearing cases of both cohorts after merging (Supplementary Fig. 6; n = 1,819 cases). The fit is P = 1.42 N^−0.929^ (slope 95% CI −0.960 to −0.898, R² = 0.650). The exponent is close to but statistically distinguishable from −1 (two-sided t = 4.44, 1,817 d.f., P = 9.3 × 10⁻⁶), so the expected number of culprits per case rises about 1.4-fold across the two orders of magnitude of panel size observed here rather than being exactly scale-invariant. Binned median residuals stay within 0.249 log₁_0_ units of zero.

### Context-injection experiment

To test whether the pair-specific structure can be exploited by naming the other causative organisms of a sample, a context arm was trained and evaluated against the baseline arm (Supplementary Fig. 7). In the context arm the other causative organisms of the same sample are added to the prompt, noised during training and supplied at inference as the first-pass predictions of the same model; both arms use identical data, decoding and scoring, and only the prompt differs. Strata are defined on the reference labels, so stratum sizes are identical in the two arms and every within-stratum comparison is paired by construction; the strata contain 261, 400, 372 and 1,346 positives for 0, 1, 2 and ≥3 other causative organisms in 2025 Q1, against headline denominators of 2,379 positives among 182,875 candidate rows and 819 culprit-bearing cases. The follower test, with its organism list fixed before the context arm was evaluated, covers *P. micra* (8 positives), *S. haemolyticus* (16), HHV-7 (65), HHV-6 (32), the *E. cloacae* complex (52) and *S. pneumoniae* (9).

### Prompt-block ablation

Each arm rewrites the evaluation set so that one block of fields carries the missing-value token that block already uses when the field is genuinely absent, rather than deleting the line, using a familiar missing-value representation. The comparison measures sensitivity to masking the available fields and may also reflect redundancy and changes in the input distribution. Row identifiers and labels are unchanged, verified row by row. The blocks are: clinical context (age, sex, immune status, underlying disease, presentation, diagnosis); sampling site; knowledge-base annotation (coloniser flag and organism description); the nine quantitative mNGS indices (species and genus specific reads, species and genus detection rank, genome coverage, depth ratio, Shannon index, species and genus relative abundance); and organism identity (species and genus names in Latin and Chinese, Gram class). All arms use the same trained checkpoint and decision rule and are evaluated on the ≤2024 held-out cohort, so that the cross-time cohort keeps its single role as the generalisation test. Per-arm metrics are plotted in Supplementary Fig. 2 and released as source data.

### Orthogonal microbiological reference

The clinical record carries, per specimen, the organisms found concordantly by culture, smear or nucleic-acid testing, together with fields recording whether culture and mNGS agreed and whether mNGS missed an organism culture had grown. Free-text organism names were normalised by stripping quantity expressions (scientific notation, copies per mL, colony counts, growth percentages) and specimen-source prefixes, splitting on the same separators the label builder uses, and applying a synonym table checked against the strings actually present in the candidate panels. Names were then resolved to candidate detection rows by exact match on any of the species, genus, Latin or parenthetical-abbreviation aliases of a row, falling back to a unique substring match. Of 295 (organism, specimen) pairs in the held-out cohort, 253 resolved (85.8%); 22 of those record a culture taken from a different specimen than the sequenced one, identified by a source prefix such as urine or ascites, and are excluded, leaving 231 pairs with same-specimen evidence (Fig. 6c). The comparison is insensitive to how strictly names are normalised, giving 0.822 to 0.840 for the read and 0.867 to 0.875 for the model across the normalisation rules we tried, and the record’s own field for organisms grown by culture but absent from mNGS is negative for every pair in this subset, which describes this selected subset and does not estimate the frequency of sequencing misses across the cohort. Confidence intervals are Wilson intervals and the paired comparison is an exact McNemar test on the discordant pairs. Per-pair counts, including the excluded pairs and the reason for each exclusion, are released as source data.

### Data handling and figure provenance

The analysed tables carry an anonymous specimen code and, where recorded, the coarse clinical fields described above. Direct identifiers were excluded from the analysis tables. Patient grouping was derived inside the hospital and supplied as an anonymous key. Figure source tables and available analysis scripts are intended to accompany the benchmark release, with provenance for values taken from study summaries.

## Data Availability

All data produced in the present study are available upon reasonable request to the authors.

## Data availability

The adjudication benchmark and the per-figure source data will be made available to the research community on publication. The released material is derived from clinical report files and consists of de-identified, row-level tables: per-organism reporting fields, the coarse clinical descriptors, the adjudication labels and an anonymous specimen code. No raw sequencing reads are included, since none were used in this work, and patient-level clinical free text is not released. Because the tables derive from clinical records, access is controlled: requests should be addressed to the corresponding authors and will be reviewed by the Department of Laboratory Medicine, Ren Ji Hospital, with a decision normally within 30 days. Access is granted for non-commercial research under a data-use agreement that prohibits re-identification and onward transfer. The specimen codes used in this paper cannot be resolved to a patient outside the hospital. A versioned data record and persistent identifier will be provided with the approved release.

Figure-level source tables are intended to accompany the release. Each table will identify the relevant unit of analysis, including specimen, organism detection, species–site combination, organism pair, module or evaluation arm, as appropriate.

Three additional source tables are planned to document quantities not displayed in full in the figures. The first gives every (case, pathogen) culprit call of the 215 blind re-adjudicated cases, labelled by which read made it and by the candidate row it resolved to, or the reason it could not be resolved; Fig. 1d plots its per-pathogen summary. The second gives both directions of every evaluable organism pair — the conditional causative rates, odds ratio, z score and q value with the focal organism taken each way — of which Fig. 5a shows one direction under a fixed orientation rule. The third gives every (organism, specimen) pair with independent microbiological confirmation, including the pairs excluded for an unresolved name or for a culture taken from a different specimen than the sequenced one, of which Fig. 6c shows the aggregate.

## Code availability

The training, evaluation and analysis code, including prompt templates, figure scripts and the script for regenerating inter-rater statistics, is planned for release on publication. A specific repository, versioned archive and licence will be provided with the release.

## Acknowledgements

We thank the staff of the Department of Laboratory Medicine, Ren Ji Hospital, for the clinical metagenomic testing whose reports this work analyses, and the clinical teams whose recorded adjudications form the labels. This work was supported by the National Key Research and Development Program of China (2025YFC3409100 to M.L.), the National Natural Science Foundation of China (W2411070 to M.L. and 82604593 to M.W.) and the Shanghai Science and Technology Innovation Action Plan Innovative Pharmaceutical and Medical Device Product Application Demonstration (24SF1903500 to M.L.). The funders had no role in study design, data analysis, the decision to publish or the preparation of the manuscript.

## Author contributions

M.W., M.L. and Z.D. designed the study. M.W., S.Z., K.X., S.L. and L.Liu assembled the clinical cohort and performed the causative-pathogen adjudication. Y.Z. and C.F. built the benchmark, trained and evaluated the models and performed the statistical analysis. Y.P. and L.L. curated the reference database and the taxonomic merging rules. Z.D. supervised the computational work and M.L. supervised the clinical work. Y.Z. and C.F. wrote the manuscript with input from all authors. All authors contributed to manuscript review.

## References

1. Wilson, M. R. et al. Clinical metagenomic sequencing for diagnosis of meningitis and encephalitis. N. Engl. J. Med. 380, 2327–2340 (2019). 10.1056/NEJMoa1803396

2. Chiu, C. Y. & Miller, S. A. Clinical metagenomics. Nat. Rev. Genet. 20, 341–355 (2019). 10.1038/s41576-019-0113-7

3. Gu, W., Miller, S. & Chiu, C. Y. Clinical metagenomic next-generation sequencing for pathogen detection. Annu. Rev. Pathol. 14, 319–338 (2019). 10.1146/annurev-pathmechdis-012418-012751

4. Blauwkamp, T. A. et al. Analytical and clinical validation of a microbial cell-free DNA sequencing test for infectious disease. Nat. Microbiol. 4, 663–674 (2019). 10.1038/s41564-018-0349-6

5. Miller, S. et al. Laboratory validation of a clinical metagenomic sequencing assay for pathogen detection in cerebrospinal fluid. Genome Res. 29, 831–842 (2019). 10.1101/gr.238170.118

6. Park, S. Y. et al. Plasma microbial cell-free DNA sequencing from over 15,000 patients identified a broad spectrum of pathogens. J. Clin. Microbiol. 61, e0185522 (2023). 10.1128/jcm.01855-22

7. Simner, P. J., Miller, S. & Carroll, K. C. Understanding the promises and hurdles of metagenomic next-generation sequencing as a diagnostic tool for infectious diseases. Clin. Infect. Dis. 66, 778–788 (2018). 10.1093/cid/cix881

8. Salter, S. J. et al. Reagent and laboratory contamination can critically impact sequence-based microbiome analyses. BMC Biol. 12, 87 (2014). 10.1186/s12915-014-0087-z

9. Chinese Thoracic Society. Consensus of clinical pathways of metagenomic next-generation sequencing test in diagnosis of lower respiratory tract infections in China. Zhonghua Jie He He Hu Xi Za Zhi 46, 322–335 (2023). 10.3760/cma.j.cn112147-20220701-00553

10. Zhou, H. Several issues in interpreting the results of metagenomic next-generation sequencing of lower respiratory tract specimens. Zhonghua Jie He He Hu Xi Za Zhi 46, 319–321 (2023). 10.3760/cma.j.cn112147-20221117-00903

11. Xu, C. et al. China expert consensus on the application of metagenomic next-generation sequencing for the etiological diagnosis of infections in patients with haematological diseases. Blood Sci. 7, e00241 (2025). 10.1097/BS9.0000000000000241

12. Naccache, S. N. et al. A cloud-compatible bioinformatics pipeline for ultrarapid pathogen identification from next-generation sequencing of clinical samples. Genome Res. 24, 1180–1192 (2014). 10.1101/gr.171934.113

13. Wood, D. E., Lu, J. & Langmead, B. Improved metagenomic analysis with Kraken 2. Genome Biol. 20, 257 (2019). 10.1186/s13059-019-1891-0

14. Davis, N. M. et al. Simple statistical identification and removal of contaminant sequences in marker-gene and metagenomics data. Microbiome 6, 226 (2018). 10.1186/s40168-018-0605-2

15. Singhal, K. et al. Large language models encode clinical knowledge. Nature 620, 172–180 (2023). 10.1038/s41586-023-06291-2

16. Thirunavukarasu, A. J. et al. Large language models in medicine. Nat. Med. 29, 1930–1940 (2023). 10.1038/s41591-023-02448-8

17. Hager, P. et al. Evaluation and mitigation of the limitations of large language models in clinical decision-making. Nat. Med. 30, 2613–2622 (2024). 10.1038/s41591-024-03097-1

18. Brodeur, P. G. et al. Performance of a large language model on the reasoning tasks of a physician. Science 392, 524–527 (2026). 10.1126/science.adz4433

19. Johnson, A. E. W. et al. MIMIC-III, a freely accessible critical care database. Sci. Data 3, 160035 (2016). 10.1038/sdata.2016.35

20. Yoon, W. et al. LCD benchmark: long clinical document benchmark on mortality prediction for language models. J. Am. Med. Inform. Assoc. 32, 285–295 (2025). 10.1093/jamia/ocae287

21. Steinig, E. et al. Large language models enable consensus-level interpretation in metagenomic diagnostics. Preprint at medRxiv (2026). 10.64898/2026.07.29.26358751

22. Li, T. et al. GPAS: an online AI system for rapid and accurate pathogen identification and LLM-based interpretation. Preprint at medRxiv (2026). 10.64898/2026.02.18.26346517

23. Yang, A. et al. Qwen3 technical report. Preprint at arXiv (2025). 10.48550/arXiv.2505.09388

24. Sati, H. et al. The WHO Bacterial Priority Pathogens List 2024: a prioritisation study to guide research, development, and public health strategies against antimicrobial resistance. Lancet Infect. Dis. 25, 1033–1043 (2025). 10.1016/S1473-3099(25)00118-5

25. Brooke, J. S. *Stenotrophomonas maltophilia*: an emerging global opportunistic pathogen. Clin. Microbiol. Rev. 25, 2–41 (2012). 10.1128/CMR.00019-11

26. Sharma, R. et al. *Acinetobacter baumannii*: a comprehensive review of global epidemiology, clinical implications, host interactions, mechanisms of antimicrobial resistance and mitigation strategies. Microb. Pathog. 204, 107605 (2025). 10.1016/j.micpath.2025.107605

27. Rath, S. et al. Pathogenic functions of host microbiota. Microbiome 6, 174 (2018). 10.1186/s40168-018-0542-0

28. Hahn, W. O. et al. Multidrug-resistant Corynebacterium striatum associated with increased use of parenteral antimicrobial drugs. Emerg. Infect. Dis. 22, 1908–1914 (2016). 10.3201/eid2211.160141

29. Wexler, H. M. Bacteroides: the good, the bad, and the nitty-gritty. Clin. Microbiol. Rev. 20, 593–621 (2007). 10.1128/CMR.00008-07

30. Otto, M. Molecular basis of Staphylococcus epidermidis infections. Semin. Immunopathol. 34, 201–214 (2012). 10.1007/s00281-011-0296-2

31. Kotton, C. N. et al. The fourth international consensus guidelines on the management of cytomegalovirus in solid organ transplantation. Transplantation 109, 1066–1110 (2025). 10.1097/TP.0000000000005374

32. Ljungman, P. et al. Definitions of cytomegalovirus infection and disease in transplant patients for use in clinical trials. Clin. Infect. Dis. 64, 87–91 (2017). 10.1093/cid/ciw668

33. Pociupany, M., Snoeck, R., Dierickx, D. & Andrei, G. Treatment of Epstein–Barr virus infection in immunocompromised patients. Biochem. Pharmacol. 225, 116270 (2024). 10.1016/j.bcp.2024.116270

34. Fishman, J. A., Gans, H. & AST Infectious Diseases Community of Practice. Pneumocystis jiroveci in solid organ transplantation: guidelines from the American Society of Transplantation Infectious Diseases Community of Practice. Clin. Transplant. 33, e13587 (2019). 10.1111/ctr.13587

35. Yu, Q. et al. Outcomes and prognostic factors of non-HIV patients with *Pneumocystis jirovecii* pneumonia and pulmonary CMV co-infection: a retrospective cohort study. BMC Infect. Dis. 17, 392 (2017). 10.1186/s12879-017-2492-8

36. Heylen, J. et al. Acute invasive pulmonary aspergillosis: clinical presentation and treatment. Semin. Respir. Crit. Care Med. 45, 69–87 (2024). 10.1055/s-0043-1777769

37. Kalil, A. C. et al. Management of adults with hospital-acquired and ventilator-associated pneumonia: 2016 clinical practice guidelines by the Infectious Diseases Society of America and the American Thoracic Society. Clin. Infect. Dis. 63, e61–e111 (2016). 10.1093/cid/ciw353

38. Antimicrobial Resistance Collaborators. Global burden of bacterial antimicrobial resistance in 2019: a systematic analysis. Lancet 399, 629–655 (2022). 10.1016/S0140-6736(21)02724-0

39. Cortese, I., Reich, D. S. & Nath, A. Progressive multifocal leukoencephalopathy and the spectrum of JC virus-related disease. Nat. Rev. Neurol. 17, 37–51 (2021). 10.1038/s41582-020-00427-y

40. Kotton, C. N. et al. The second international consensus guidelines on the management of BK polyomavirus in kidney transplantation. Transplantation 108, 1834–1866 (2024). 10.1097/TP.0000000000004976

41. Jain, S. et al. Community-acquired pneumonia requiring hospitalization among U.S. adults. N. Engl. J. Med. 373, 415–427 (2015). 10.1056/NEJMoa1500245

42. Gadsby, N. J. & Musher, D. M. The microbial etiology of community-acquired pneumonia in adults: from classical bacteriology to host transcriptional signatures. Clin. Microbiol. Rev. 35, e0001522 (2022). 10.1128/cmr.00015-22

43. Bartlett, J. G. Anaerobic bacterial infection of the lung. Anaerobe 18, 235–239 (2012). 10.1016/j.anaerobe.2011.12.004

44. Kobo, O. et al. The pyogenic potential of the different Streptococcus anginosus group bacterial species: retrospective cohort study. Epidemiol. Infect. 145, 3065–3069 (2017). 10.1017/S0950268817001807

45. Plummer, E. L., et al. Are Mycoplasma hominis, Ureaplasma urealyticum and Ureaplasma parvum associated with specific genital symptoms and clinical signs in nonpregnant women? Clin. Infect. Dis. 73, 659–668 (2021). 10.1093/cid/ciab061

46. Tay, W. H., Chong, K. K. L. & Kline, K. A. Polymicrobial-host interactions during infection. J. Mol. Biol. 428, 3355–3371 (2016). 10.1016/j.jmb.2016.05.006

47. Murray, J. L., Connell, J. L., Stacy, A., Turner, K. H. & Whiteley, M. Mechanisms of synergy in polymicrobial infections. J. Microbiol. 52, 188–199 (2014). 10.1007/s12275-014-4067-3

48. Cliff, N. Dominance statistics: ordinal analyses to answer ordinal questions. Psychol. Bull. 114, 494–509 (1993). 10.1037/0033-2909.114.3.494

49. Benjamini, Y. & Hochberg, Y. Controlling the false discovery rate: a practical and powerful approach to multiple testing. J. R. Stat. Soc. B 57, 289–300 (1995). 10.1111/j.2517-6161.1995.tb02031.x

50. Kruskal, W. H. & Wallis, W. A. Use of ranks in one-criterion variance analysis. J. Am. Stat. Assoc. 47, 583–621 (1952). 10.1080/01621459.1952.10483441

51. Newman, M. E. J. & Girvan, M. Finding and evaluating community structure in networks. *Phys*. Rev. E 69, 026113 (2004). 10.1103/PhysRevE.69.026113

52. Blondel, V. D., Guillaume, J.-L., Lambiotte, R. & Lefebvre, E. Fast unfolding of communities in large networks. J. Stat. Mech. 2008, P10008 (2008). 10.1088/1742-5468/2008/10/P10008

53. Hubert, L. & Arabie, P. Comparing partitions. J. Classif. 2, 193–218 (1985). 10.1007/BF01908075

54. Mantel, N. & Haenszel, W. Statistical aspects of the analysis of data from retrospective studies of disease. J. Natl Cancer Inst. 22, 719–748 (1959). 10.1093/jnci/22.4.719

55. Kirkwood, J. G. Statistical mechanics of fluid mixtures. J. Chem. Phys. 3, 300–313 (1935). 10.1063/1.1749657

