## Supplementary Information (Figures 1-7) for "Clinical pathogen adjudication in metagenomic sequencing using a large language model"

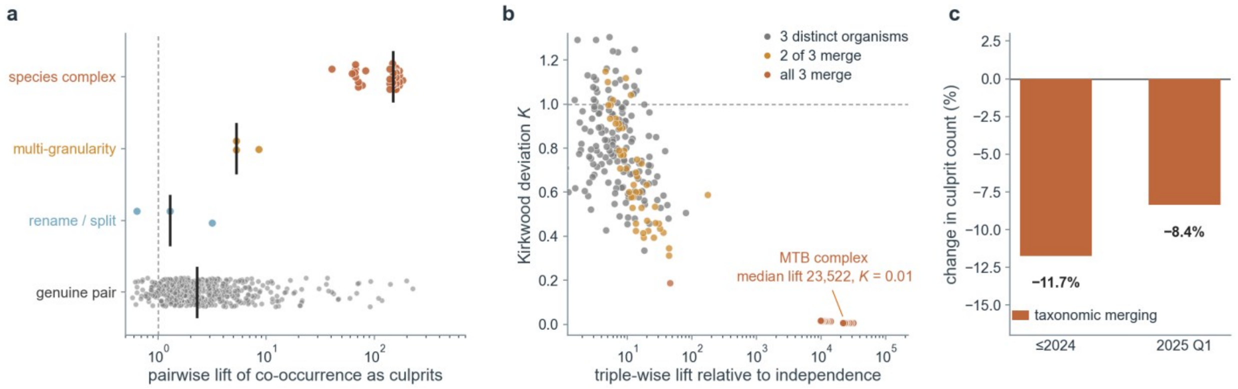


### Supplementary Figure 1 Effects of taxonomic reporting artefacts on coadjudication counts

a, Pairwise lift for every co-culprit pair with ≥3 co-culprit events and ≥5 culprit calls per member (n = 544 pairs), by pair class. Point, one pair (vertically jittered); black bar, class median; dashed line, lift = 1. Genuine biological pairs, n = 500, median lift 2.28; rename/split candidates, n = 3, median 1.28; multi-granularity reporting, n = 3, lift 5.3–8.6; species complexes, n = 38, median 148, up to 183. b, Third-order structure for every triple with ≥6 co-culprit events (n = 297 triples). x, triple-wise lift relative to independence; y, the Kirkwood third-order deviation K, K = 1 meaning the triple adds nothing beyond the pairwise structure. Colour separates triples in which all three members merge into one entity (n = 85, median lift 23,522, median K = 0.01), two of three merge (n = 48) and three distinct organisms (n = 164, median K = 0.81). c, Effect of taxonomic merging on the number of culprits, by cohort, as a percentage of the originally reported culprit rows (2,738 and 2,379). Bold figure, net change: −11.7% (≤2024) and −8.4% (2025 Q1). Neither cohort contains paired DNA and RNA libraries, so the whole of the change is taxonomic.

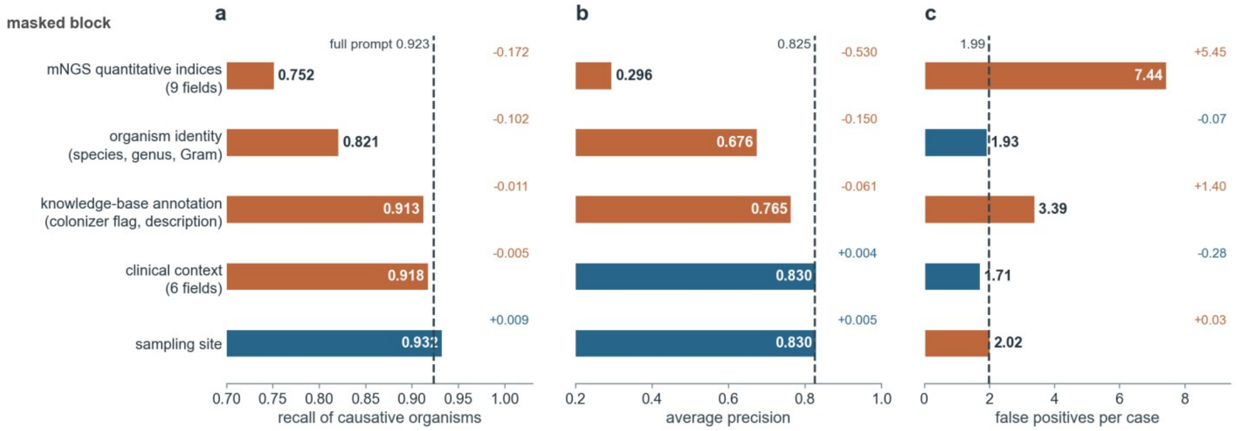


### Supplementary Figure 2 Model sensitivity to masking prompt fields

The same checkpoint and evaluation rows were used with one prompt block masked per arm (Methods). a, Recall of culprits. b, Average precision. c, False positives per case. Masking the nine quantitative mNGS indices costs 0.172 of recall, takes average precision from 0.825 to 0.296 and raises false positives per case from 1.99 to 7.44; masking organism identity costs 0.102 of recall while leaving false positives essentially unchanged; masking the knowledge-base annotation raises false positives to 3.39 per case with recall almost unchanged; masking the six clinical fields or the sampling-site label moves recall by −0.005 and +0.009. The small effect of site masking may reflect redundant site information in the candidate panel and does not establish site-independence (Fig. 3).

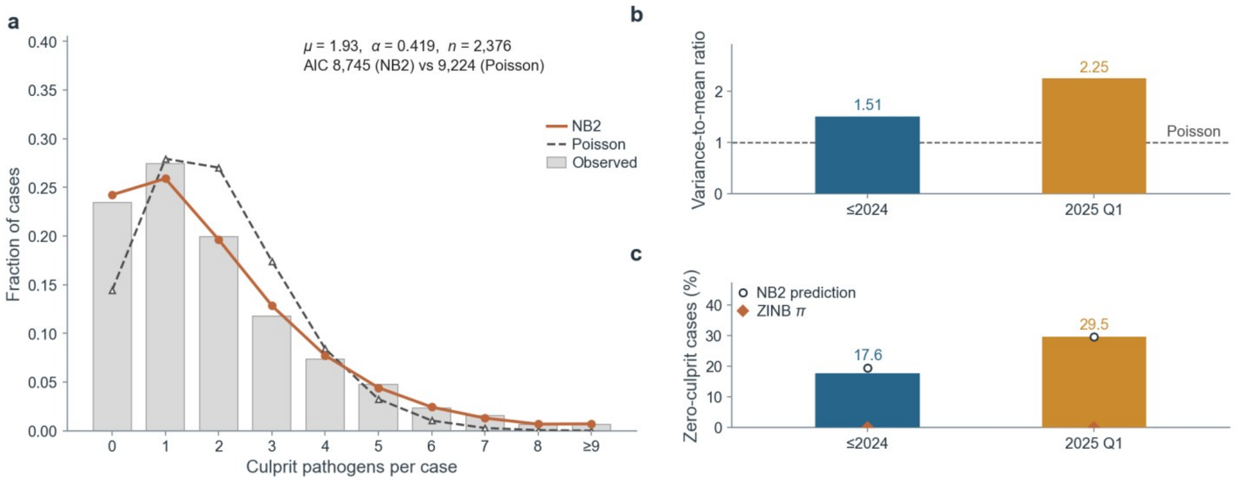


### Supplementary Figure 3 Distribution of the number of adjudicated culprit pathogens per specimen

a, Distribution of the number of culprit pathogens per case, both cohorts pooled (n = 2,376 cases; the last category collects nine or more). Grey bars, observed fractions; vermilion line with filled circles, fitted negative binomial with quadratic variance (NB2, μ = 1.93, α = 0.419, fitted variance 3.50 against an observed 3.60); grey dashed line with open triangles, Poisson with the same mean. The Poisson is rejected ($\chi^2 = 677.4$, 6 d.f., $P = 4.6 \times 10^{-143}$) while the NB2 is not ($\chi^2 = 7.5$, 8 d.f., $P = 0.48$), and the NB2 is favoured by 479 AIC units. b, Variance-to-mean ratio of the culprit count within each cohort: 1.51 (≤2024) and 2.25 (2025 Q1); dashed line at 1, the value a Poisson process would give. c, Share of cases with no culprit, by cohort: bars, observed 17.6% (≤2024) and 29.5% (2025 Q1); open circles, the shares predicted by that cohort's own NB2 fit, which reproduce them without any additional zero component; vermilion diamonds, the fitted zero-inflation weight π of a zero-inflated negative binomial, which converges to 0 in both cohorts and in the pooled fit. The fitted negative-binomial models describe both observed zero fractions; this does not establish that the underlying clinical processes are identical. Goodness-of-fit bins are the counts 0, 1, 2, … with the upper tail merged until every expected count reaches 5.

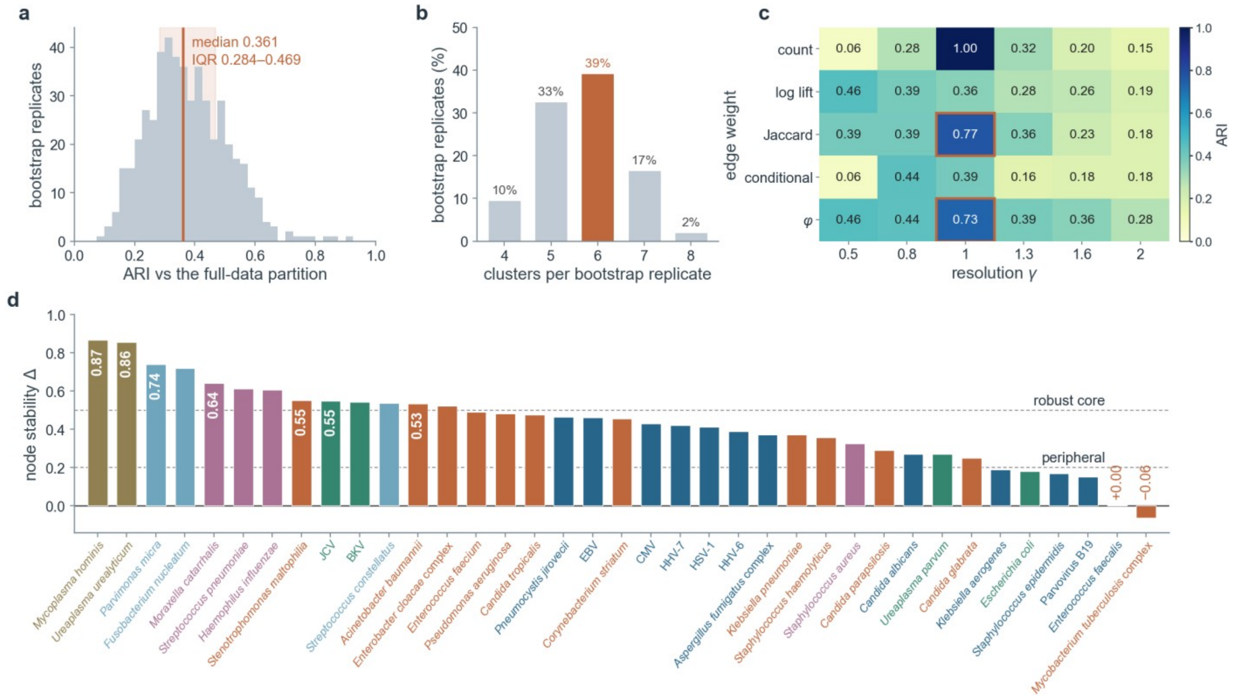


### Supplementary Figure 4 Stability of the species level coadjudication partition

a, Adjusted Rand index (ARI) between the partition from each of B = 500 case-level bootstrap replicates and the full-data partition. Vertical line, median (0.361); shaded band, interquartile range (0.284–0.469). b, Number of clusters recovered per replicate, as a percentage of replicates. Six clusters (highlighted) is the mode but accounts for only 39.2% of replicates; 4, 5, 7 and 8 clusters account for 9.6%, 32.6%, 16.6% and 2.0%. c, Sensitivity of the partition to edge weight and Louvain resolution γ. Cells give the ARI against the reference six-module partition, which is the raw-count partition at γ = 1, so that cell is 1.00 by construction. The two normalised weightings recover essentially the same communities at γ = 1 (Jaccard 0.77, φ 0.73; boxed), so the reported partition does not depend on the edge weight. d, Node stability Δ for the 37 of 49 nodes carrying an estimate, ranked (Methods; same 500 partitions as in a). Bar colour, reference module, as in Figs. 4 and 5; dashed lines at Δ = 0.5 and 0.2 delimit the robust core and the peripheral nodes; the seven organisms of the robust core carry their Δ inside their bar (0.53–0.87). The two labelled organisms at or below zero, the Mycobacterium tuberculosis complex (Δ = −0.06) and Enterococcus faecalis (Δ = +0.00), had uncertain module assignment; their module colour in Fig. 4c reflects the partition as detected and is not evidence of membership. Modules 4–6 are almost disconnected from the main graph, so their high stability is structural rather than evidential (14, 8 and 6 clustered cases).

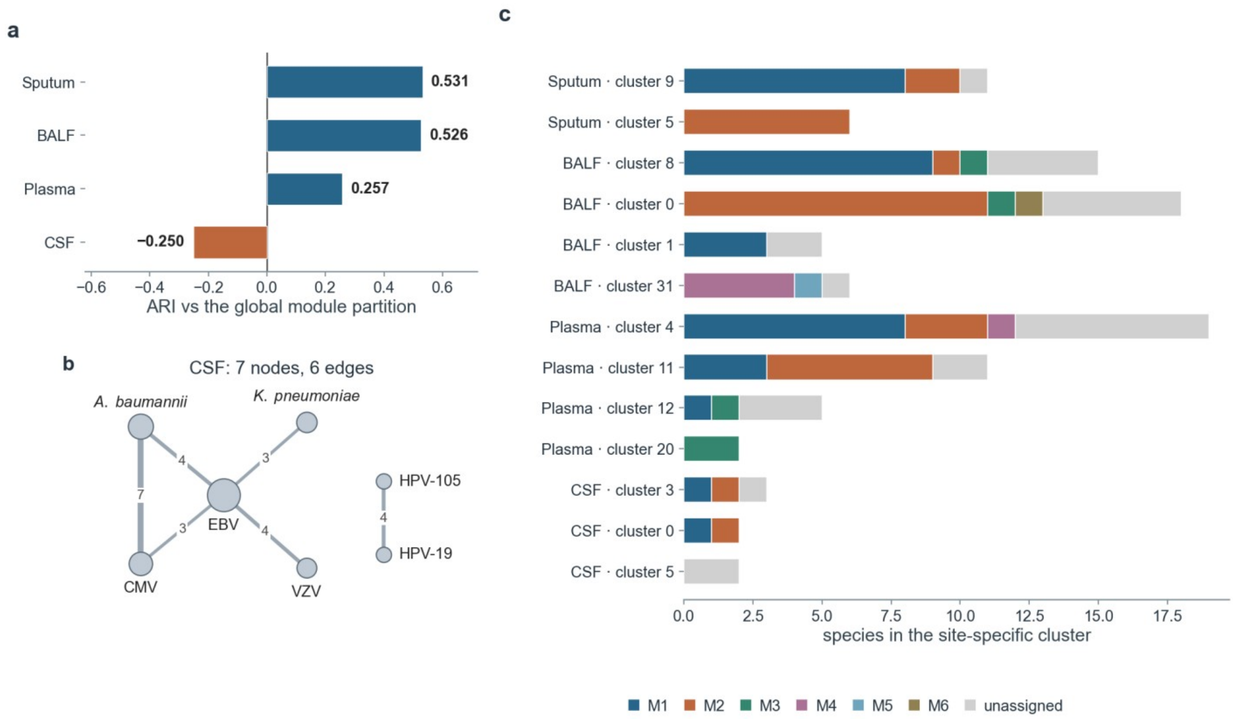


### Supplementary Figure 5 Site specific coadjudication networks

a, Agreement between the module partition learned within each site and the global partition, as the adjusted Rand index. Bars coloured by sign (blue, positive; vermilion, negative); sample, node and edge counts per site are in Methods. Sputum (0.531) and bronchoalveolar lavage fluid (0.526) recover the global structure; plasma agrees only weakly (0.257), the two largest modules there merging into one cluster, a descriptive difference requiring further evaluation; cerebrospinal fluid gives a negative index (−0.250), with interpretation limited by the sparse CSF network. b, The complete cerebrospinal-fluid co-culprit graph: 7 nodes and 6 edges from 268 samples. Node area, culprit calls for that organism in CSF; edge width and printed number, samples in which the two organisms are co-culprits. The graph is too sparse to establish a reproducible community structure. c, Composition of each site-specific cluster in terms of the six reference modules. Bar, one cluster from one site; segment length, member species drawn from each reference module, or from none (‘unassigned’). Clusters supported by fewer than 4 clustered cases are omitted; the supporting case counts are in Methods. Sputum and BALF recover modules 1 and 2 as separate clusters; in plasma the largest cluster mixes them; CSF yields only two- to three-member clusters.

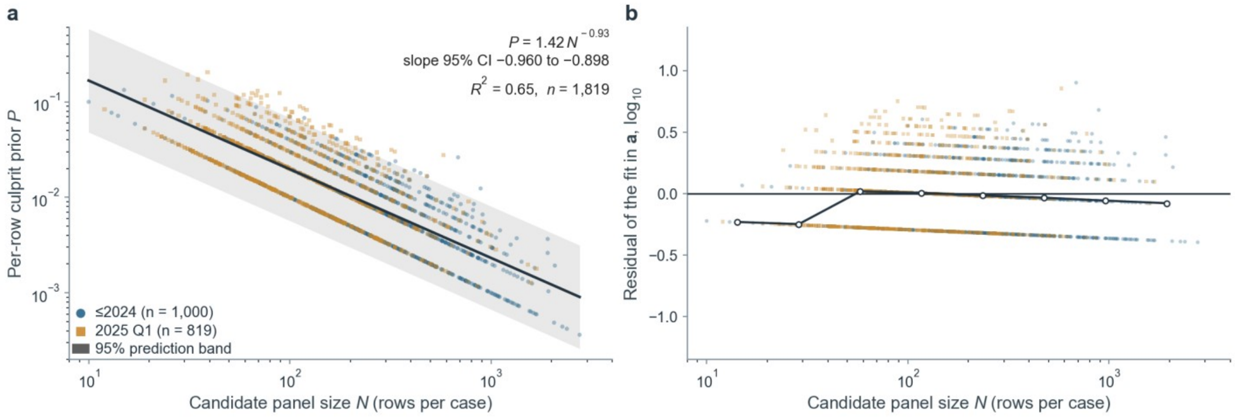


### Supplementary Figure 6 Association between candidate panel size and the proportion of culprit records

a, Per-row culprit prior P = K/N against candidate panel size N for the 1,819 cases with at least one culprit (≤2024, n = 1,000; 2025 Q1, n = 819), on log–log axes; colour and marker shape denote cohort. K, number of culprit pathogens; N, candidate rows reported for that case. Black line, ordinary least-squares fit of $\log_{10}P$ on $\log_{10}N$, $P = 1.42\ N^{-0.929}$, slope 95% CI −0.960 to −0.898, $R^2$ = 0.650; grey envelope, its 95% prediction band for a single new case. The exponent is close to, but statistically distinguishable from, −1 (two-sided t = 4.44, 1,817 d.f., $P = 9.3 \times 10^{-6}$), so the expected number of culprits per case grows slowly with panel size rather than being exactly scale-invariant. The diagonal bands in the cloud are loci of constant K, an arithmetic consequence of P = K/N at small integer K. b, Residuals of that fit, on a $\log_{10}$ scale, against N for the same cases. Horizontal line, zero residual; black circles joined by a line, medians of eight logarithmically spaced N bins containing more than five cases each. The binned medians stay within 0.249 $\log_{10}$ units of zero across two orders of magnitude of panel size, the deviation being confined to the two smallest bins (N < 41), where P = K/N is dominated by the discreteness of a small integer K.

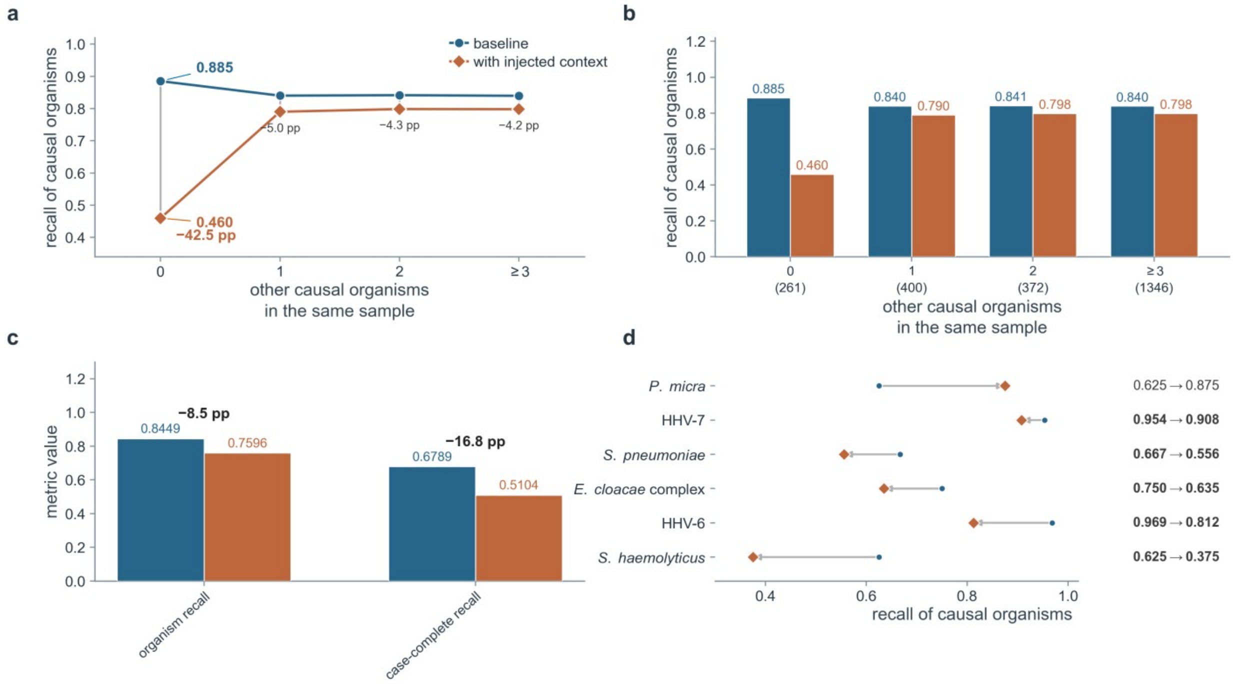


### Supplementary Figure 7 Performance after adding predicted culprit context

The baseline and context arms use identical evaluation rows; stratum definitions are based on reference labels (Methods). a, Recall of culprits on the 2025 Q1 cohort, stratified by the number of other culprits in the same sample. Circles, baseline; diamonds, context arm; grey connectors join the arms within a stratum; the change in percentage points is printed below each stratum. In the stratum with no other reference-labelled culprit recall falls from 0.885 to 0.460 (−42.5 pp, n = 261 positives); the three strata with other reference-labelled culprits change by only −5.0, −4.3 and −4.2 pp. b, Per-stratum recall of the two arms as absolute values rather than as a paired change, with stratum sizes on the axis. c, Headline metrics before and after context injection, each value printed above its bar; organism-level recall falls from 0.8449 to 0.7596 and case-level complete recall from 0.6789 to 0.5104, both arms scored on the same 182,875 candidate rows and 819 culprit-bearing cases. d, Prespecified organism subset comprising the six followers identified in Fig. 5b. Circles, baseline recall; diamonds, recall with injected context; arrows, direction of change; both recalls are printed at the right of each row. Recall of the *Enterobacter cloacae* complex falls from 0.750 to 0.635. Reference-label strata do not necessarily identify the contents of the first-pass predicted context.
